# First-In-Human Trial of Encapsulated Cell-Based Protein Producers for Localized IL-2 in Patients with High-Grade Serous Ovarian Carcinoma

**DOI:** 10.1101/2025.11.07.25339137

**Authors:** Helen D. Clark, Samira Aghlara-Fotovat, Jake Schladenhauffen, Jonathon Debonis, Juan Carlos Amador-Molina, Amanda Nash, Manish Jain, Ryan Newman, Lauren Jansen, Karen Andreas, Kelly Rangel, Travis T. Sims, Bryan Fellman, Claudio Dansky Ullman, Oladapo Yeku, Amy J. Bregar, Andrew M. Blakely, Cara Mathews, Oleg A. Igoshin, Cara Haymaker, Jose Oberholzer, Peter Rios, Daisy Lopez, Hafsa Nasir, Ira Joshi, Rima Chakrabarti, Omid Veiseh, Amir A. Jazaeri, Shannon N. Westin

**Affiliations:** Department of Gynecologic Oncology and Reproductive Medicine, The University of Texas MD Anderson Cancer Center; Houston, Texas, USA; Sentinel BioTherapeutics Inc., Houston, Texas USA; Rice Biotechnology Launch Pad, Rice University, Houston, Texas USA; Rice University, Department of Bioengineering, Houston, Texas USA; Department of Translational Molecular Pathology, The University of Texas MD Anderson Cancer Center; Houston, Texas, USA; Avenge Biotherapeutics Inc., Boston, Massachusetts, USA; Department of Hematology and Oncology, Massachusetts General Hospital, Boston, Massachusetts, USA; Center for Cancer Research, National Cancer Institute, Bethesda, Maryland, USA; Department of Obstetrics and Gynecology, Women and Infants Hospital, Providence, Rhode Island, USA; CellTrans, Inc., Chicago, IL, USA; Department of Visceral and Transplant Surgery, University of Zurich, Switzerland

**Author notes:** Co-first authors (Helen D. Clark, Samira Aghlara-Fotovat). Co-primary investigators (Omid Veiseh, Amir A. Jazaeri, Shannon N. Westin). Corresponding author: Shannon N. Westin < > 1155 Pressler St, Unit 1362, Houston, TX 77030. **Declaration of interests**: The following patents are related to this work: WO 2021/026484, WO 2021/163242, WO 2023/069993, WO 2023/070000. The registered IND number is 28550. R. Chakrabarti is a shareholder and member of the board of directors of Sentinel BioTherapeutics C. Haymaker reports research funding to institution from Sanofi, Avenge, Iovance, KSQ, Theolytics, BTG, Novartis, Astrazeneca, EMD Serono, Takeda, Obsidian, Genentech, BMS, Summit Therapeutics, Artidis and Immunogenesis and personal fees from Regeneron and stock options from Briacell outside the submitted work. O.A. Igoshin reports consulting for Avenge Bio. A.A. Jazaeri reports grants/contracts to institution from Merck, Iovance, Eli Lilly, Macrogenics, Immatics, Natera, Imunon, Outpace Bio, and Theolytics. He sevrves as an advisor for Macrogenics, Theolytics, Iovance, Immatics, Sentinel Bio, IQVIA, Premiere Research, Sentinel Bio, Duracyte, and RBL. He additionally reports consulting for GLG and Guidepoint. I. Joshi is a shareholder of CellTrans, Inc. D. Lopez is an employee of CellTrans, Inc. C. Mathews reports grants/contracts to institution from Deciphera, Astellas Pharma, Seagen, Genmab, EMD Serono, Merck, Regeneron, Moderna, Astra Zeneca, AvengeBio, Zentalis, GlaskoSmithKline, Novocure, Immunogen, Elucida, Roche/Genentech, Pfizer, Laekna, Artios, Volastra, AADI, ProfoundBio, Context Therapeutics, and REPARE. J. Oberholzer is a founder, employee, shareholder, and board member of CellTrans, Inc. P.D. Rios is an employee, shareholder, and board member of CellTrans, Inc. J. Schladenhauffen reports prior employment by Avenge Bio. He additionally reports consulting for Sentinel Bio. T.T. Sims reports grants/contracts to institution from Merck and the NIH. He additionally reports consulting for Merck and AstraZeneca. O. Veiseh reports grants/contracts to the institution from Avenge Bio (now named Sentinel Bio). He serves as a co-founder and equity holder in Duracyte Bio, Steer Bio, Sentinel Bio, and RBL. He additionally reports consulting for GLG and Guidepoint. S.N. Westin reports grants/contracts to institution from Astra Zeneca, AvengeBio, Bayer, Bio-Path, Clovis Oncology/Pharma, Daiichi Sankyo, GSK, Jazz Pharmaceuticals, Loxo, Mereo, Novartis, Nuvectis, Pfizer, Roche/Genentech, Verastem, Zentalis. She additionally reports consulting for AstraZeneca, Bayer, Caris, Clovis Oncology/Pharma&, Corcept, Daiichi Sankyo, Eisai, Genmab, Gilead, GSK, Immunocore, ImmunoGen, Incyte, Lilly, Loxo, Merck, Mereo, Mersana, NGM Bio, Nuvectis, Pfizer, Roche/Genentech, SeaGen, Verastem, Zentalis, and ZielBio. **Statement of Translational Relevance**: In this phase I dose escalation trial, we demonstrate the clinical activity of a novel drug (AVB-001) delivered intraperitoneally for the treatment of platinum-resistant high-grade serous ovarian cancer. In a prior publication, we demonstrated the preclinical efficacy of IL-2-secreting encapsulated cell therapies in mouse models. This study advances the concept into clinic and describes the first-in-human use of living IL-2 factories designed for localized intraperitoneal immunotherapy in patients with platinum-resistant ovarian cancer. This encapsulated cell technology permits sustained and self-limited delivery of local cytokine therapy, a treatment option that has previously had limited use due to challenges with administration and toxicity. This trial provides support for the safety, feasibility, and preliminary immunological activity of this approach and offers a compelling step towards a new modality of immunotherapy.

## Abstract

**Background:** Platinum-resistant high-grade serous ovarian carcinomas (HGSOC) are associated with poor therapeutic outcomes. While HGSOC frequently metastasizes to the intraperitoneal (IP) cavity, the success of IP cytokine therapies, such as IL-2, has been hampered by local toxicity and administration difficulties. AVB-001 is a novel IL-2 delivery system consisting of encapsulated, allogeneic cells engineered for constitutive human IL-2 expression.

**Methods:** This is a phase I dose-escalation trial of AVB-001 for the treatment of HGSOC (NCT05538624). A single dose of AVB-001 was administered IP laparoscopically, enabling hIL-2 doses from 0.6 to 3.6 μg hIL-2/kg/day. Safety was evaluated using NCI CTCAE v5.0. Efficacy was assessed via RECIST v1.1 criteria.

**Findings:** The trial enrolled 14 patients across four dose levels. Three patients (21.4%) experienced grade 3 treatment-related adverse events (TRAEs); no grade 4-5 TRAEs were reported. There was one unconfirmed partial response lasting 29 days (ORR 7.1%). Stable disease was observed in seven patients, with a median duration of 2.57 months (range 2.03-4.23). Pharmacokinetics demonstrated dose-dependent increases in serum IL-2, peaking at 1 day post-implantation. Immunological analyses revealed sustained CD8+ and CD4+ T-cell proliferation without corresponding proliferation in regulatory T cells. Dose-dependent CTLA-4 receptor upregulation was observed on CD8+ and CD4+ T cells, whereas PD-1 and TIM-3, remained unchanged.

**Conclusion:** In patients with HGSOC, AVB-001 is safe and effectively activates cytotoxic T cells, supporting further investigation of this locoregional immunotherapy.

Graphical Abstract

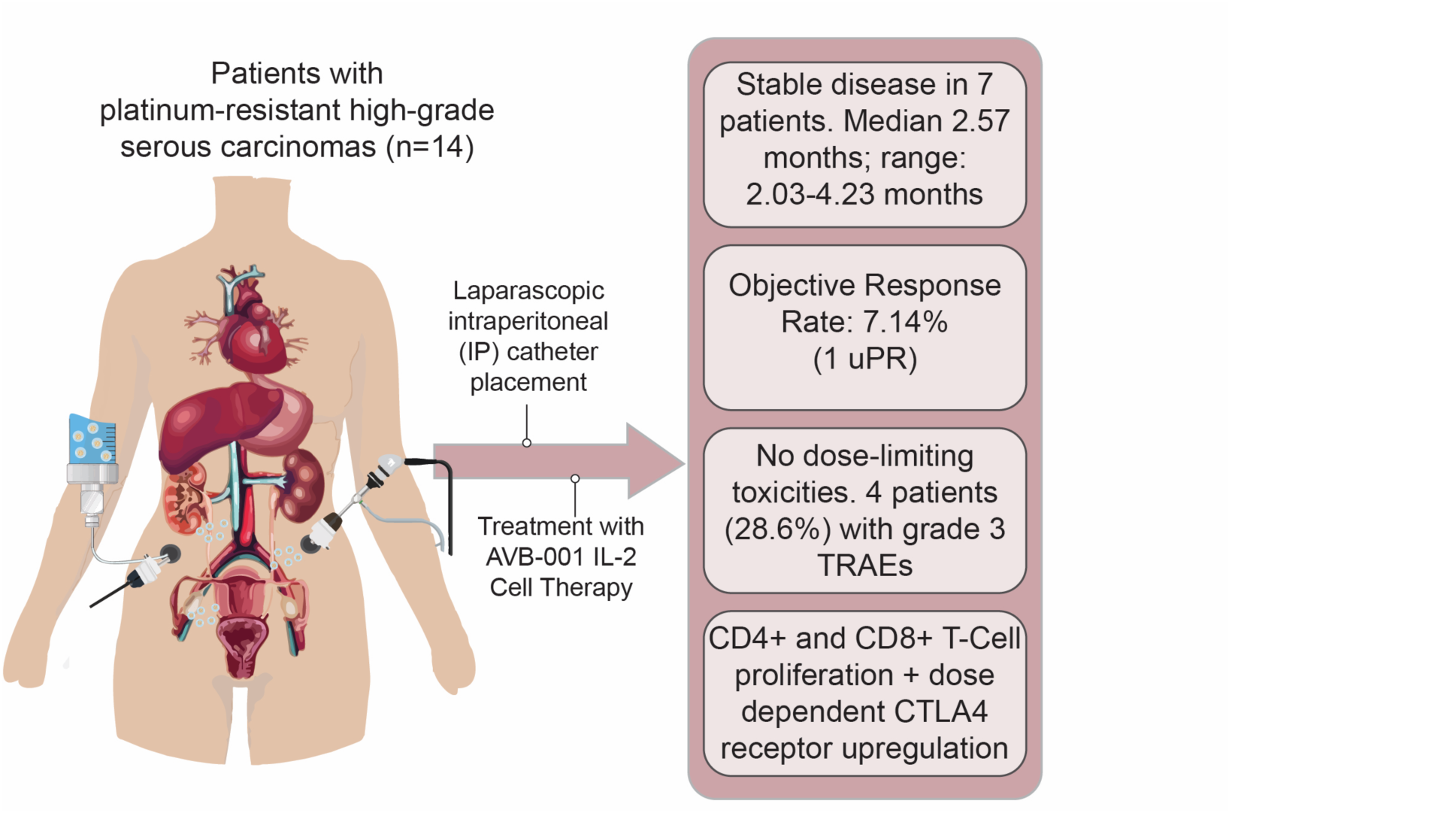

## INTRODUCTION

Cancers originating from the ovary, fallopian tube, or peritoneum account for over 20,000 new cancer cases annually in the United States and over 12,000 deaths per year^1^. Of these cases, the majority are of high-grade serous histology, hereafter collectively referred to as high-grade serous ovarian carcinomas (HGSOC), which are often diagnosed at an advanced stage and are characterized by an aggressive natural history and a high rate of recurrence^2^. While approximately 70-80% of patients respond to frontline treatment with platinum-doublet chemotherapy and surgical cytoreduction, the majority ultimately experience recurrence^3,4^. In the recurrent setting, the disease becomes incurable and response rates decline with subsequent lines of therapy, resulting in the inevitable development of platinum-resistant disease with an associated median overall survival (OS) of approximately 12 months^5^.

A common feature of HGSOC is its propensity to metastasize throughout the peritoneal cavity^6^. This behavior has generated particular interest in the development of efficacious locoregional treatments administered via an intraperitoneal (IP) route^7,89^. The IP route of administration is intriguing because it permits a several-fold increase in drug concentration within the abdominal cavity and less systemic toxicity compared to the intravenous (IV) route^10–13^. For advanced-stage surgically debulked ovarian cancer patients, IP chemotherapy has previously been associated with a potential survival advantage^14,15^. Widespread adaptation, however, has not occurred due to administration difficulties and the potential for local toxicity^16,17^.

Immunotherapy has previously demonstrated promise as a strategy for treating ovarian cancer. In particular, IV administration of interleukin-2 (IL-2), a cytokine known for its ability to activate and expand cytotoxic T cells and natural killer (NK) cells, has demonstrated significant activity in the treatment of melanoma and renal cell carcinoma^18,19^. However, its short serum half-life has required it to be administered in high doses with an elevated risk of severe inflammatory toxicities^20,21^. To mitigate these side effects, researchers have investigated localized intraperitoneal (IP) delivery of IL-2. In a pivotal 1997 study, Edwards et al. reported an overall response rate (ORR) of 25.7% in PROC patients receiving IP IL-2^22^. A subsequent trial in 2009 yielded comparable results, with an ORR of 25%; however, 45% of patients were unable to complete treatment due to adverse events, primarily catheter-associated complications^23^. While these studies underscore the therapeutic potential of locally delivered IL-2, they also highlight the urgent need for safer, more effective delivery strategies.

This challenge led to the development of a bioengineered, encapsulated cell therapy platform that enables localized, continuous secretion of biologic agents, including IL-2. Preclinical studies have demonstrated that these IL-2–encapsulated protein producers (ePPs) achieve sustained, high local cytokine concentrations with minimal systemic distribution in both murine and non-human primates^13,24^. In murine models of ovarian cancer, colorectal cancer, and malignant pleural mesothelioma, IL-2 cytokine factories induced robust activation and proliferation of cytotoxic T cells within the tumor microenvironment, leading to complete tumor regression and the establishment of durable immunological memory upon re-challenge^13,24^.

This study describes the first-in-human administration of localized IL-2 producing ePPs (termed AVB-001) to the IP space in patients with platinum-resistant HGSOC. The primary objectives were to evaluate the feasibility, safety, and tolerability of AVB-001 after a single IP administration and to determine the maximum tolerated dose (MTD) and the recommended phase II dose (RP2D). Translational objectives were to characterize the pharmacokinetics and immunological pharmacodynamics of AVB-001.

## MATERIALS AND METHODS

### Capsule Manufacturing

#### Alginate Preparation

GMP-grade SLG20 sodium alginate was hydrated with sterile 0.9 % saline at a concentration of 1.4 % w/v and mixed overnight under agitation. The alginate was then filtered through a 0.2 um PES filter and aseptically aliquoted. Aliquots were stored at 2-8C until use.

#### Capsule Cell Viability

In triplicate, 10 capsules were incubated at 37C for 30 minutes in 200 uL of DMEM/F12 supplemented to contain 1 mg/mL alginate lyase, 10% v/v FBS (Cytiva), and 5 mM EDTA. Following incubation, capsules were mechanically disrupted by gentle pipetting. For research cell counts and viability, cell suspensions were mixed 1:1 with Trypan Blue and added to microfluid cell counting slides. Slides were immediately analyzed on a Countess (Thermofisher) automated cell counting to quantify cell concentration and viability. Clinical samples were instead added to a vial 1-casette and analyzed on an NC-200 (Chemometec).

#### Quantification of hIL-2 production in capsules

In triplicate, 5 capsules were plated in a single well of a 48-well plate and incubating in DMEM/F12 culture media supplemented to 10% v/.v FBS media for 4-24 hours. The supernatant was collected and frozen at –20C. Samples were analyzed for hIL-2 concentration using a Human IL-2 Quantikine ELISA Kit (R&D Systems)

#### Mycoplasma Testing-Drug Product

A sample of formulated drug product storage solution collected. Sample submitted to an external vendor (IDEXX Bioanalytic, Columbia MO) for mycoplasma testing via real-time PCR.

#### Mycoplasma Testing-Drug Substance

Approximately 5e5-1e6 cells were aliquoted in a tube and frozen at –80C. Sample submitted to an external vendor (IDEXX Bioanalytic, Columbia MO) for mycoplasma testing via real-time PCR.

#### Sterility Testing

A sample of formulated drug product storage solution collected and submitted to an external vendor (IDEXX Bioanalytic, Columbia MO). Sample was inoculated into growth media and observed for signs of contamination. For clinical drug product, samples were prepared similarly and submitted to (Eurofins, Pennsylvania). Sample was tested according to USP <71> standards. Samples were similarly collected for rapid sterility testing used for generation of a COT. Samples from drug product were collected, inoculated into BACT/ALERT media vials, and sent to an external vendor (Eurofins, Pennsylvania) for growth monitoring.

#### Endotoxin Testing

The storage solution of formulated test and control article was collected and submitted to Associates of Cape Cod, Falmouth MA or Focus Labs, Florida for research and clinical samples respectively. Samples were assessed for kinetic-chromogenic endotoxin detections. LOD for clinical samples was <0.05 EU/mL.

#### Residual Barium Testing

The storage solution of formulated test and control article was collected and submitted to an external vendor (Intertek, Whitehouse NJ or Eurofins, Pennsylvania for research and clinical samples respectively) for Ion Chromatography-Mass Spectrometry analysis. For clinical samples the LOD was 6.25 ppm

#### Capsule Fracture Strength

Twenty individual capsules were analyzed for initial fracture using a TA-XT. Plus texture analyzer (TA Instruments, Delaware). Capsules were compressed to 90% strain from which the applied force at the initial fracture force was determined.

#### Residual Bovine Serum Albumin Testing

A sample of drug product storage solution was collected in low-protein binding microcentrifuge tubes. Samples were run on BSA ELISA kit (Cygnus Technologies, North Carolina).

### Study Design

This was an open-label, multicenter, phase I dose escalation study of AVB-001 in patients with platinum-resistant, high-grade serous adenocarcinoma of the ovary, primary peritoneum, or fallopian tube (NCT05538624). Patients in the study were required to have histologically confirmed, metastatic or unresectable, platinum-resistant, high-grade serous adenocarcinoma. Platinum-resistance was defined as either progression during initial platinum-based chemotherapy, relapse within 6 months of receiving initial platinum-containing therapy, or progression after receiving at least 2 lines of platinum-containing chemotherapy. Because of the route of administration, all patients were required to have intraperitoneal disease with evidence of measurable disease on computed tomography (CT) or magnetic resonance imaging (MRI). There was no preferred site identified for catheter placement and capsule administration, rather the catheter was inserted as per standard of care considerations with laparoscopy.

The primary objectives for the dose escalation phase were to evaluate the feasibility, safety, and tolerability of AVB-001 after a single IP administration and to determine the maximum tolerated dose (MTD) and the recommended phase II dose (RP2D). The secondary objectives included translational correlates to assess the pharmacokinetics and immunologic response in the peripheral blood. In addition, preliminary efficacy was evaluated using the Response Evaluation Criteria in Solid Tumors (RECIST) v1.1.

The dose escalation phase consisted of a single dose administration of 1 of 4 dose levels of AVB-001. The starting dose was 0.6 μg hIL-2/kg/day. The maximum dose was capped at 3.6 μg hIL-2/kg/day. The study drug was administered into the IP cavity via a minimally invasive laparoscopy procedure. A Bayesian optimal interval (BOIN) 3+3 design was utilized to identify the MTD/RP2D with a targeted toxicity rate for the MTD of approximately 30%. Patients were enrolled and treated in cohorts of 3 patients per cohort. The first cohort of 3 patients was staggered to minimize potential risks associated with the first-in-human administration of AVB-001. Escalation to the next higher dose level was based on the targeted toxicity rate for the MTD of approximately 30%. The decision to escalate or de-escalate per BOIN design was based on whether the observed probability of a DLT at the current dose is ≤0.236 or ≥0.359, respectively. If the probability of DLT is at or below 0.236, dose escalation to the next higher dose occurred. If the probability of DLT is >0.236 and <0.359, the dose was maintained for the next 3 patients.

DLTs were evaluated for a 4 week period from time of study drug administration. Hematologic DLTs were defined as grade 4 neutropenia lasting more than 7 days, febrile neutropenia, grade 4 anemia not resolving in 7 days, grade 4 thrombocytpenia, or grade 3 thrombocytopenia lasting more than 7 days. Non-hematologic DLTs were defined as grade 3 or higher toxicities probably or possibly related to study procedures, grade 3 or higher toxicities involving major organs systems, grade 3 or higher cytokine release syndrome, grade 2 or higher neurotoxicity that does not resolve to grade 1 or lower within 72 hours, AST/ALT elevation 3 times the upper limit of normal with concurrent increase in bilirubing to 2 times the upper limit of normal, AST/ALT elevation over 8 times the upper limit of normal, or total bilirubin over 5 times the upper limit of normal. Escalation to the next dosing level was permitted when the probability of a DLT was equal to or less than 23.6%. The trial was initially designed to include a phase II dose expansion phase; however, the study was ultimately terminated after the dose escalation phase due to funding limitations.

Patients were recruited via physician referral and word of mouth. Eligible participants were required to have histologically confirmed, metastatic or unresectable, platinum-resistant high-grade serous adenocarcinoma (HGSC) originating in the ovary, fallopian tube, or peritoneum. Patients may have received a maximum of 5 lines of prior therapy, have an ECOG score of 0-1, and must have evidence of measurable disease as defined by RECIST v1.1. Key exclusion criteria included those with non-serous or mixed histologies, individuals with a synchronous primary or prior malignancy within 3 years, prior administration of IL-2 or IL-2-like molecules, or any prior IP gene-targeted therapy.

This study involves human participants. This study was approved by the US Food and Drug Administration and the Institutional Review Boards (IRB) at the University of Texas MD Anderson Cancer Center (IRB-approved protocol #2022-0420), Massachusetts General Hospital, the Center for Cancer Research at the National Cancer Institute, and Women and Infants Hospital. All participants provided written informed consent and all procedures were conducted in accordance with the International Council for Harmonisation guidelines for Good Clinical Practice.

### Study Assessments

Response to treatment within the study cohort was assessed with serial imaging via CT or MRI. Imaging assessment was conducted at screening and at post-treatment assessment intervals. Serial imaging was continued until progression was noted. For patients with imaging demonstrating progression of disease, an additional confirmatory assessment was conducted after 4 weeks due to the potential for pseudo-progression with response to immunotherapy. For efficacy assessment, patients must have received the full dose of the study drug and undergone at least one post-baseline efficacy assessment.

Adverse event assessment was conducted continuously from the time of the administration of the study drug.. Nature, incidence, relatedness, and severity of all adverse events, including the occurrence of any dose-limiting toxicities (DLTs), were reported according to the National Cancer Institute (NCI) Common Terminology Criteria for Adverse Events (CTCAE) v5.0. All patients exposed to AVB-001 were evaluated for safety.

To allow for assessment of pharmacodynamics and to determine the potential for AVB-001 to induce immunogenicity, collection of peripheral blood was performed at baseline and at specified post-treatment timepoints.

### Flow Cytometry Sample Prep and Staining

Aliquots of peripheral blood mononuclear cells (PBMCs) and IP fluid samples were stored in liquid nitrogen for retrospective analysis. Upon thawing, samples underwent cell count and cell viability quality control checks on a Cellometer K2 Cell Counter (Nexcelom Bioscience). Samples with no viable cells were excluded from analysis. PBMCs were spun down, washed, and resuspended in FACS wash buffer (1X DPBS with 1% BSA). Flow cytometry staining was performed for myeloid (**Supplementary Table S5**) and function and memory T cells (**Supplementary Table S6**) respectively.

Following staining, acquisition was performed on a LSRFortessa TM X-20 (Becton Dickinson, New Jersey) flow cytometer using the FACSDiva software. Samples failing to meet the quality control metric of >100 CD45^+^ live cell events were excluded from analysis. Acquired data was subsequently procced on FlowJo v10 software.

#### Myeloid Panel Gating

Lymphocytes were identified based on size and granularity using a forward scatter area (FSC-A) vs. side scatter area (SSC-A) plot. Next, single cells were identified by excluding doublets using FSC-Height (FSC-H) vs. FSC-A and sequential SSC-Height (SSC-H) vs. SSC-A plots. A time gate was applied to ensure consistent flow and eliminate irregularities. Live cells were then identified using viability dye. CD45-positive events, representing leukocytes, were selected using an FSC vs. CD45 plot. A dump gate was applied to remove unwanted cells by gating HLA-DR expression vs dump gate which excluded CD3/CD19/CD20/CD55 positive cells. Monocytes and dendritic cells were gated by the expression of HLA-DR vs CD14. HLA-DR is a marker of antigen-presenting cells and CD14 is a monocyte marker. From the HLA-DR+ CD14+ population, cells were further gated on CD16 vs CD14 expression. CD14+ cells were classified into three monocyte subsets based on CD16 expression: classical monocytes (CD14+ CD16-), intermediate monocytes (CD14+ CD16+), and non-classical monocytes (CD14dim CD16+). The HLA-DR-CD14+ population was further gated by CD33 expression. From the HLA-DR+ CD14-population, CD11b vs CD11c gating was used to distinguish activated monocytes and dendritic cells (CD11b-CD11c+). Further gating of the CD11b+ CD11c+ population by CD141 expression vs CD1c expression was preformed to identify DC1 (CD141+) and DC3 (CD141-) subsets. The CD11b+ CD11c-population was further gated by CD123 used to identify plasmacytoid dendritic cells (pDCs).

#### Functional and Memory T Cell Gating

Lymphocytes were identified based on size and granularity using a forward scatter area (FSC-A) vs side scatter area (SSC-A) plot. Single cells were identified by gating for FSC-Height (FSC-H) vs FSC-A and sequential SSC-Height (SSC-H) vs. SSC-A. A time gate was applied to ensure consistent flow and eliminate irregularities. Live cells were then identified using viability dye. CD45-positive events, representing leukocytes, were selected using an FSC vs. CD45 plot. The CD45+ population was then gated by CD3 expression to select T cells, which were subsequentially gated by CD4+ and CD8+ populations. Memory subsets were defined based on CCR7 and CD45RA expression, distinguishing naïve (CCR7+CD45RA+), central memory (CCR7+CD45RA−), effector memory (CCR7−CD45RA−), and terminal effector (CCR7−CD45RA+) cells. Effector memory cells were classified into 4 subsets based on CD27 and CD28 expression: EM1 (CD27+, CD29+), EM2 (CD27+, CD28-, seen only in CD8+ population), EM2 (CD27-, CD28-), and EM4 (CD27-, CD28+). The EM2 was only seen in CD8+ population. Functional analysis follows, with markers like ICOS, CD25, CD28, and Ki67 identifying activation and proliferation states, while PD-1, Tim3, and CTLA-4 assess exhaustion. FoxP3 vs CD25 gating was used to identify regulatory T cells from the CD4+ population.

### Clinical Serum Analysis

Serum cytokine concentrations were measured by MEDPACE Reference Laboratories. Serum IL-2 levels were measured by MEDPACE Bioanalytical Laboratories via electrochemiluminescent assay. All samples were processed, shipped, and stored according to the laboratory’s specified requirements.

### Mathematical Modeling of AVB-001 Pharmacokinetics

A mathematical model of AVB-001 pharmacokinetics was developed based on a previously published model of the system ^13^. Model parameters were estimated with nonlinear mixed effects modeling with proportional error using the nlmixr package in R with the SAEM parameter estimation method ^35,36^. Initial parameter estimates were obtained by estimating individual patient parameter sets with the naïve pooled approach, and all parameters were assumed to be log-normally distributed. Fixed and random effect parameters were determined by first assuming all parameters are random effects, then removing random effects on all parameters with a shrinkage value greater than 30%. The final version of the model was then used to estimate the amount of AVB-001-produced IL-2 in the peritoneal cavity throughout the course of treatment.

### Animal Model and Study Design

Four Mauritian Cynomolgus macaques, adult females (10–15 years old, 2–8 kg) and adult males (5–7 years old, 3–9 kg) were used in this study. All macaque study subjects were naïve to prior procedures. Each subject was assigned a unique 4-digit numerical identifier (CN9171, CN9313, CN9317, CN9314). They were housed with ad libitum access to food and water under standard light/dark cycles. Cage checks were conducted throughout the study to monitor behavior and overall health. All studies were approved by the Institutional Animal Care and Use Committee (IACUC).

Capsule delivery was performed under standard sterile precautions in a surgical suite. The anterior abdomen was shaved and prepped from the xyphoid process to the pubis. A small (2 cm) supraumbilical incision was made, and a 5–12 mm trocar was inserted. Pneumoperitoneum was created using CO₂ at a pressure of 10–14 mmHg._The peritoneal cavity was warmed with heated saline and a laparoscopic camera was introduced through the trocar to provide visualization. Under laparoscopic guidance, a second small incision (1 cm) was made, and a 5–12 mm trocar was inserted. Capsules were delivered into the peritoneal cavity using an Argyle Tenckhoff Peritoneal catheter (Ref 8810889003) connected to a 60 mL sterile Luer lock syringe. Capsules were suspended in sterile saline at a ratio of approximately 7 mL of saline per mL of capsule, with no more than 5 mL of capsule delivered per syringe. Each primate received a dose of approximately 10ug/kg/day AVB-001 hIL-2. Control primates received saline only, with volume calculated based on weight. Laparoscopic videos and images were recorded to confirm the absence of abnormalities within the peritoneal cavity. Upon completion of the procedure, the small incisions were closed in layers using Vicryl 3-0 for muscle and a 4-0 subcuticular suture for the skin.

Animals were monitored throughout the study for behavioral or health abnormalities. Clinical observations for signs such as labored breathing, facial redness, vomiting, tremors, lethargy, or diarrhea were conducted three times weekly, with abnormal findings documented. Body weight and rectal temperature were measured thoughout the study. Blood samples were collected via intravenous draws for hematological, clinical chemistry, and coagulation analyses.

### Statistical Analysis

Descriptive summary statistics were employed to describe demographics and clinical characteristics of the evaluable patients. Flow cytometry data were normalized using the range function in JMP Pro 15 (Min-Max approach), adjusting values to a range of 0 to 1. Normalization was performed separately for each set of surface checkpoint inhibitor markers. Data was then transformed to visualize fold differences relative to the mean of the baseline points for each checkpoint inhibitor. The peak value from each patient was selected. All data were analyzed and visualized using GraphPad Prism (version 10.2.2, GraphPad Software, San Diego, CA, USA). Statistical significance was assessed using one-way analysis of variance (ANOVA) with Tukey’s post hoc test for multiple comparisons. A family-wise alpha threshold of 0.05 was applied, corresponding to a 95% confidence interval (CI). Data are presented as mean ± SEM, and statistical significance is denoted in the figure where applicable.

### Data Availability

All requests must be submitted in writing to the corresponding author via email. The consent form signed by the study participants did not include consent for the sharing of raw data. Therefore, the raw data cannot be made available. Any additional information required to reanalyze the data reported in this paper, as well as the study protocol and statistical analysis plan, is available from the lead contact upon request.

## RESULTS

### Clinical Grade (GMP) Manufacturing of AVB-001 for Human Testing

AVB-001 is an encapsulated cell product consisting of allogeneic, genetically enhanced retinal pigment epithelial (RPE) cells engineered to secrete human IL-2 (hIL-2). The manufacturing process follows a standardized workflow involving cell expansion, batch harvesting, and encapsulation via electrostatic spraying, a technique validated across multiple preclinical models, including murine and non-human primate studies ^13,24^. Cell expansion occurs over 28 days, yielding approximately 1.2 billion cells before sequential harvesting and encapsulation (D29) (**Figure 1**). Encapsulation is achieved using a syringe pump-driven electrostatic spraying system, where alginate-embedded cells are formed into uniform capsules with a controlled 1.5 mm diameter. Following encapsulation, pooled batches undergo stringent quality control (QC) assessments, with each batch formulated and stored for up to 10 days prior to preparation for clinical administration.

**Figure 1.**
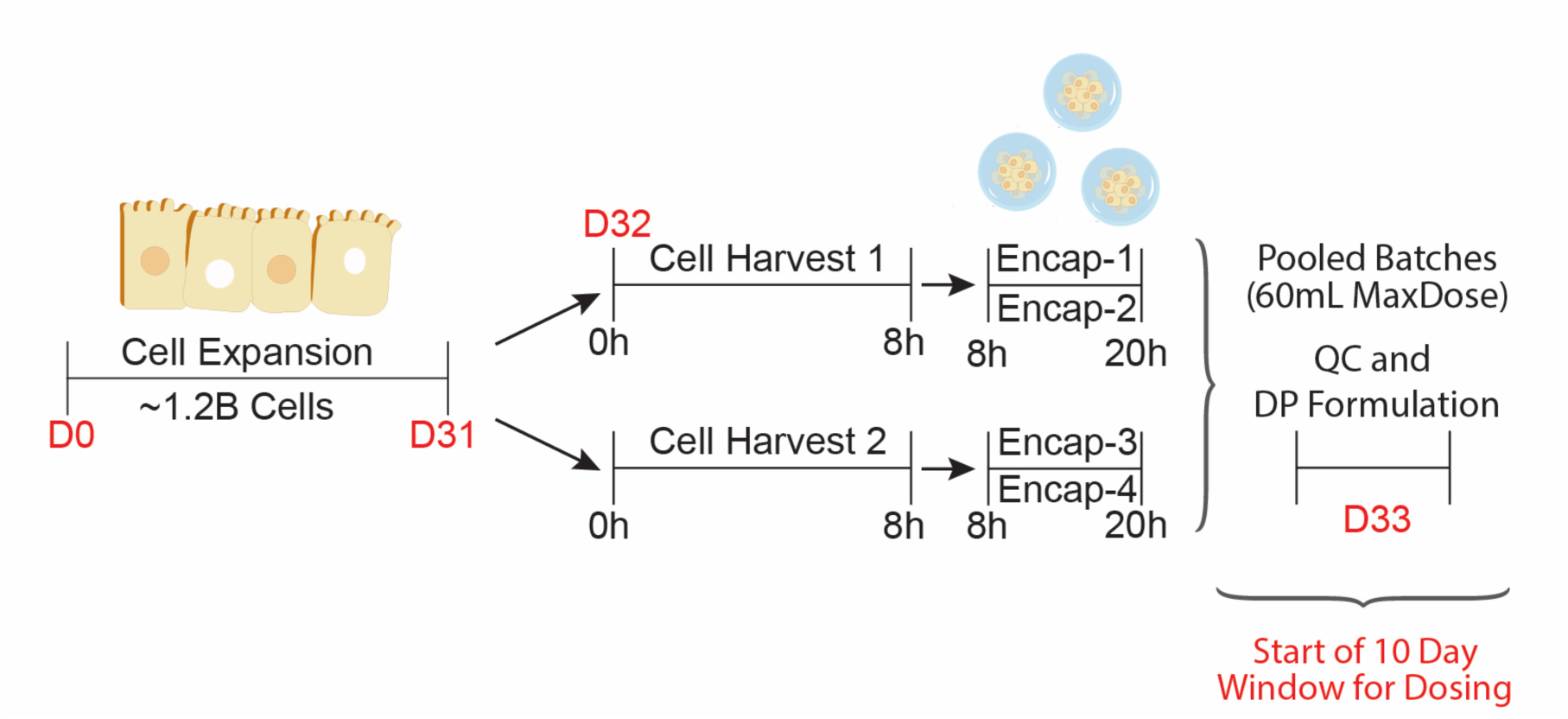
Manufacturing timeline for drug product development. Schematic illustration showing RPE cell expansion, two parallel batched harvests generating two encapsulation runs each, and the corresponding timelines.

To ensure clinical-grade reproducibility, rigorous manufacturing criteria were evaluated across all production runs, demonstrating consistent adherence to predefined specifications (**Table 1**). Key drug substance attributes, including cell viability (>90%, measured at 96 ± 1%) and IL-2 secretion (>0.5 pg/cell/day, measured at 0.8 ± 0.2 pg/cell/day), met or exceeded acceptance criteria. Drug product assessments confirmed stable pH (7.19 ± 0.06), osmolality (302 ± 2 mOsm/kg), and capsule morphology (>70% conforming, achieved at 97 ± 5%). Furthermore, each batch was evaluated for sterility, endotoxin levels, and residual fetal bovine serum (FBS), with all tested samples meeting predefined safety thresholds. Notably, the encapsulated product exhibited robust fracture strength (>15.0 g, measured at 18.9 ± 1.9 g), ensuring mechanical stability throughout handling and administration (**Table 1**). The consistency of these results underscores the robustness and reproducibility of AVB-001 manufacturing, supporting its clinical translation and scalable production for future studies.

**Table 1.**
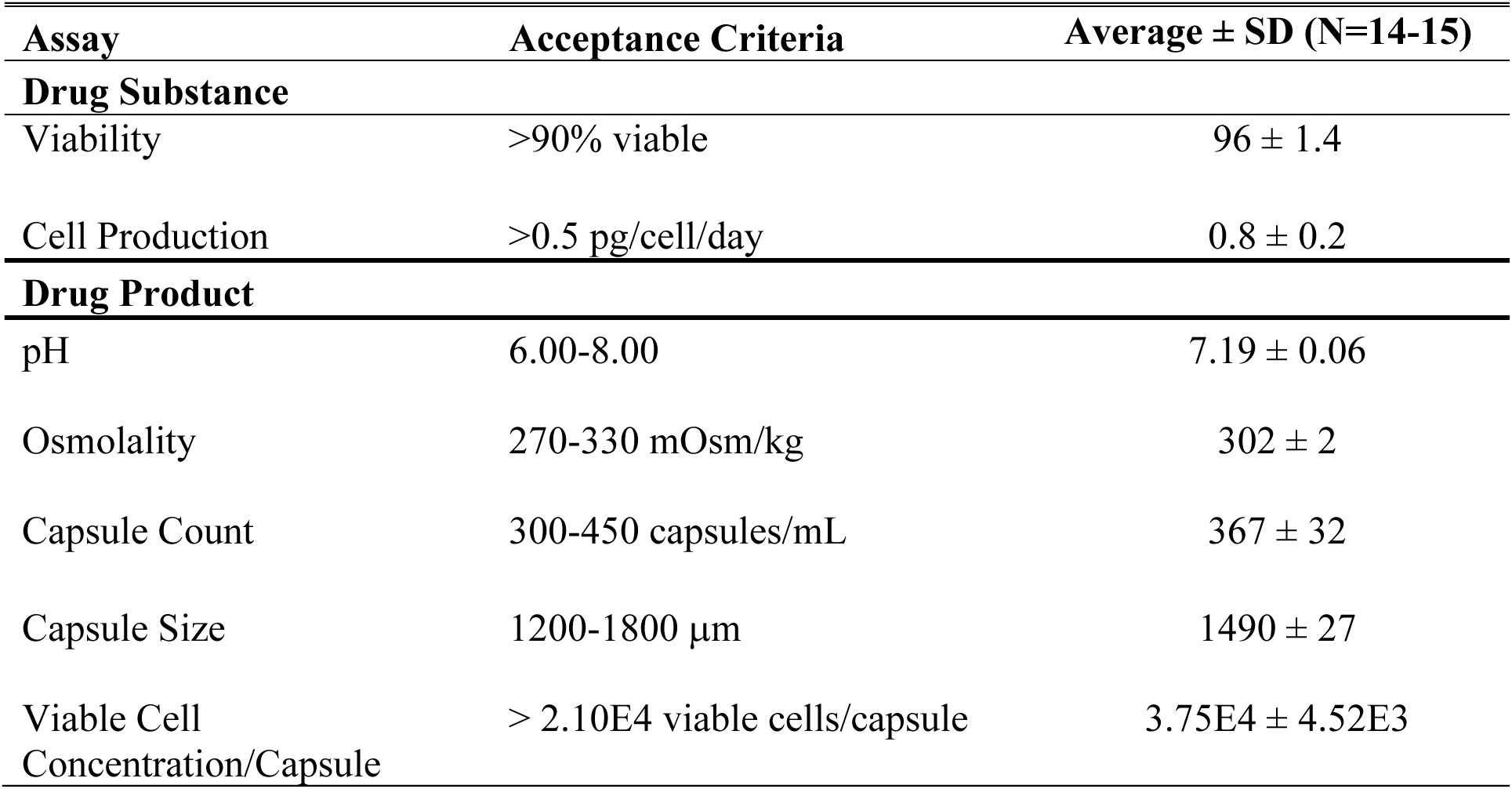
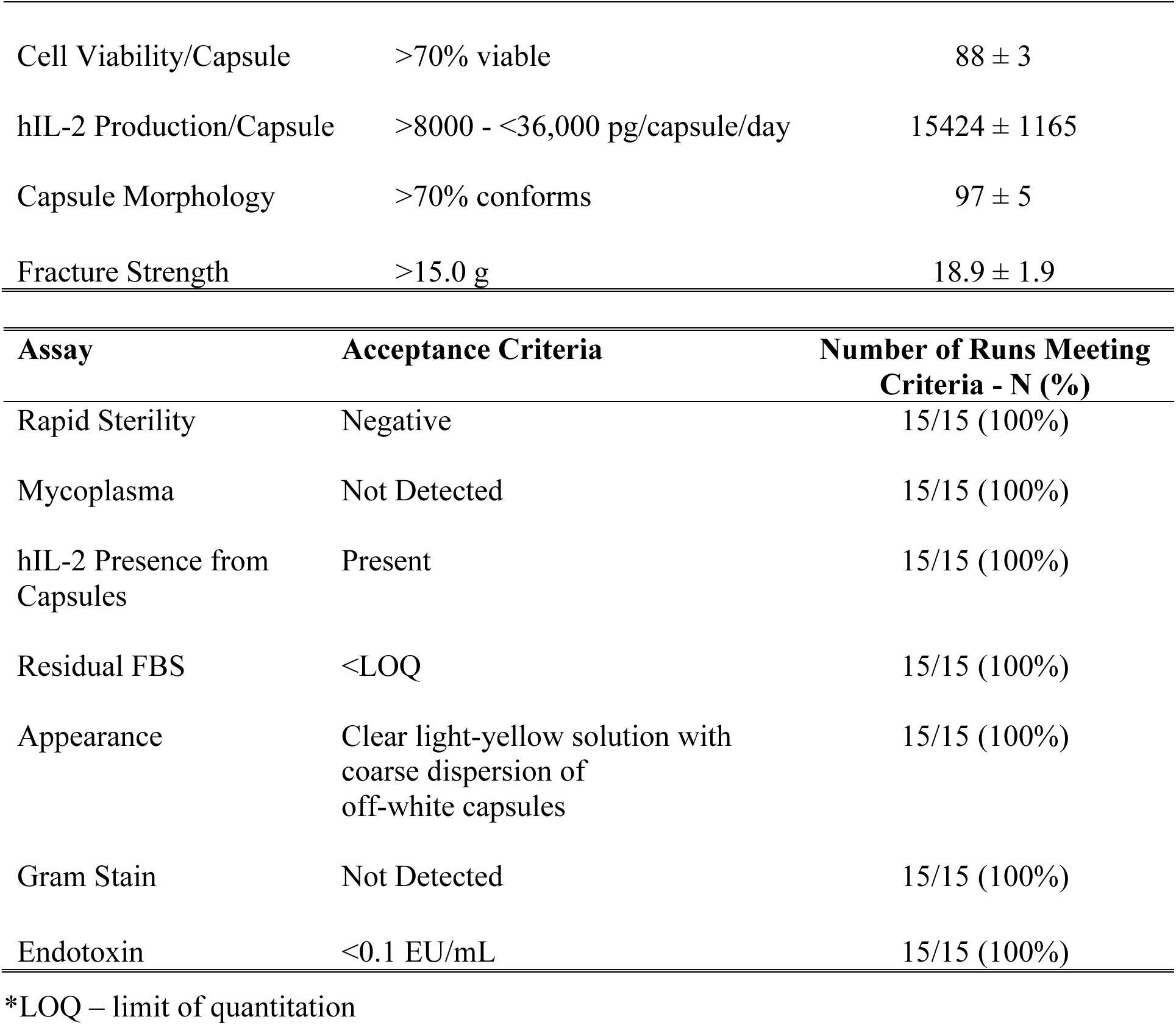
Manufacturing criteria. Drug product manufacturing criteria, acceptance criteria, and average values measured per patient dose.

### Patient Characteristics

The trial enrolled 14 patients between November 2022 and November 2023. Eleven patients had primary ovarian cancer (78.6%), two had primary fallopian tube cancer (14.2%), and one had primary peritoneal cancer (7.1%). All patients were evaluable for efficacy and safety. Clinical and demographic information for the study population is depicted in **Table 2**. Of note, five patients had previously received immunotherapy (35.7%), eleven had previously received bevacizumab (78.6%), and the median number of prior lines of therapy was 2 (range 1-4). Three patients received dose level 1, three received dose level 2, five received dose level 3, and three received dose level 4, respectively.

**Table 2.** Clinical and demographic characteristics of the study population.

| Characteristic | AVB-001 (n=14) |
| --- | --- |
| Age (in years at enrollment) |  |
| Median (min-max) | 68 (47-75) |
| Cancer Type (N, %) |  |
| Ovarian | 11 (78.6) |
| Fallopian | 2 (14.2) |
| Primary Peritoneum | 1 (7.1) |
| Baseline CA125 Level (N,%) |  |
| Normal (<38) | 3 (21.4) |
| High (>38) | 11 (78.6) |
| Race (N, %) |  |
| White/Caucasian | 14 (100) |
| Black | 0 (0) |
| Asian | 0 (0) |
| Other | 0 (0) |
| Ethnicity (N, %) |  |
| Hispanic | 1 (7.1) |
| Non-Hispanic | 13 (92.9) |
| Baseline ECOG score (N, %) |  |
| 0 | 8 (57.1) |
| 1 | 6 (42.9) |
| Prior Therapy (N, %) |  |
| Median prior lines (range) | 2 (1-4) |
| PARP-inhibitor | 7 (50) |
| Immunotherapy | 5* (35.7) |
| Bevacizumab | 11 (78.6) |

\* Two patients were previously enrolled in randomized trials that involved immune checkpoint inhibition versus placebo. Of the remaining three patients, two received immune checkpoint inhibitors and one received a therapeutic cancer vaccine.

### Safety

There was one adverse event initially noted as a dose-limiting toxicity (DLT) that occurred at dose level 3 in the setting of bowel perforation. This dose level was expanded without further DLTs noted, and thus dose escalation was again permitted. There were 11 patients (78.6%) who experienced any treatment-related adverse event. The most common side effects were anemia, chills, eosinophilia, fatigue, fever, and rash, as shown in **Table 3**. There were no grade 4 or 5 treatment-related adverse events; however, 3 patients (21.4%) did experience a grade 3 treatment-related adverse event. One patient experienced bowel perforation, two episodes of anemia, and fatigue. The other two patients experienced nausea and rash, respectively. The MTD and RP2D for AVB-001 were not reached.

**Table 3.** Treatment-related adverse events.

| Adverse Event | No. (%) of patients |  |  |
| --- | --- | --- | --- |
|  | Grade 1 | Grade 2 | Grade 3 |
| Abdominal Discomfort | 1 (7.1) | 0 (0) | 0 (0) |
| Abdominal Distention | 1 (7.1) | 0 (0) | 0 (0) |
| Abdominal Pain | 0 (0) | 2 (14.3) | 0 (0) |
| Alk Phos Increased | 0 (0) | 1 (7.1) | 0 (0) |
| ALT Elevated | 3 (21.4) | 0 (0) | 0 (0) |
| Anemia | 2 (14.3) | 0 (0) | 1 (7.1) |
| Anorexia | 1 (7.1) | 0 (0) | 0 (0) |
| Ascites | 1 (7.1) | 1 (7.1) | 0 (0) |
| AST Elevated | 3 (21.4) | 0 (0) | 0 (0) |
| Bloating | 0 (0) | 1 (7.1) | 0 (0) |
| Bowel Perforation | 0 (0) | 0 (0) | 1 (7.1) |
| Bullous Dermatitis | 0 (0) | 1 (7.1) | 0 (0) |
| Capillary Leak Syndrome | 0 (0) | 1 (7.1) | 0 (0) |
| Chills | 3 (21.4) | 1 (7.1) | 0 (0) |
| Diarrhea | 1 (7.1) | 2 (14.3) | 0 (0) |
| Dyspepsia | 0 (0) | 1 (7.1) | 0 (0) |
| Edema | 0 (0) | 1 (7.1) | 0 (0) |
| Eosinophilia | 5 (35.7) | 0 (0) | 0 (0) |
| Fatigue | 3 (21.4) | 2 (14.3) | 1 (7.1) |
| Fever | 3 (21.4) | 0 (0) | 0 (0) |
| Gas Pain | 1 (7.1) | 0 (0) | 0 (0) |
| GGT Increased | 0 (0) | 1 (7.1) | 0 (0) |
| Hypokalemia | 1 (7.1) | 0 (0) | 0 (0) |
| Hypomagnesemia | 1 (7.1) | 0 (0) | 0 (0) |
| Hyponatremia | 0 (0) | 1 (7.1) | 0 (0) |
| Hypotension | 1 (7.1) | 0 (0) | 0 (0) |
| Myalgias | 0 (0) | 1 (7.1) | 0 (0) |
| Nausea | 1 (7.1) | 0 (0) | 1 (7.1) |
| Pallor | 1 (7.1) | 0 (0) | 0 (0) |
| Platelet Count Decreased | 2 (14.3) | 0 (0) | 0 (0) |
| Pleural Effusion | 1 (7.1) | 0 (0) | 0 (0) |
| Pruritus | 0 (0) | 1 (7.1) | 0 (0) |
| Rash | 1 (7.1) | 3 (21.4) | 1 (7.1) |

### Efficacy

Objective tumor response, assessed according to RECIST v1.1, demonstrated an unconfirmed partial response (uPR) in one patient (ORR: 1/14, 7.1%), with duration of response lasting 29 days. An additional seven patients (50%) experienced stable disease (SD), with a median duration of clinical stability lasting 2.57 months (range: 2.03–4.23) (**Figure 2**). A dose-dependent trend in disease stabilization was observed, with the highest dose cohort (3.6 μg hIL-2/kg/day) exhibiting the longest mean duration of disease control. The percentage change from baseline in the target lesion size for each patient is illustrated in **Figure 3**. The best response as a percentage change from baseline tumor volume is shown in **Supplemental Figure 1**.

**Fig. 2.**
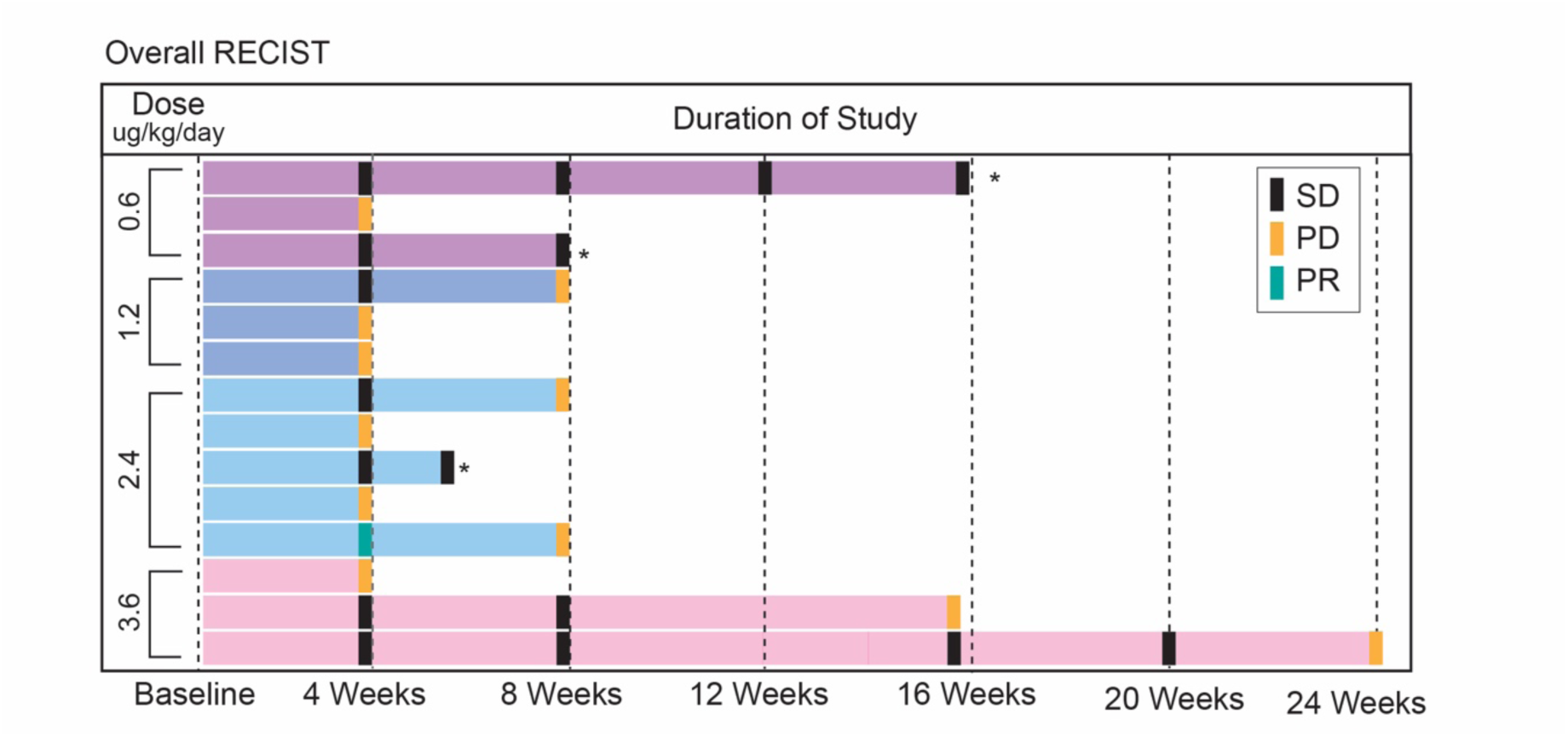
Swimmer plot Disease status (Stable Disease [SD], Progressive Disease [PD], Unconfirmed Partial Response [uPR]) by cohort based on response evaluation criteria in solid tumors (RECIST v.1.1) for the evaluable study population (n=14) stratified by dose level. Each bar represents one patient in the study. Asterisks indicate clinical progression without corresponding radiologic progression.

**Fig. 3.**
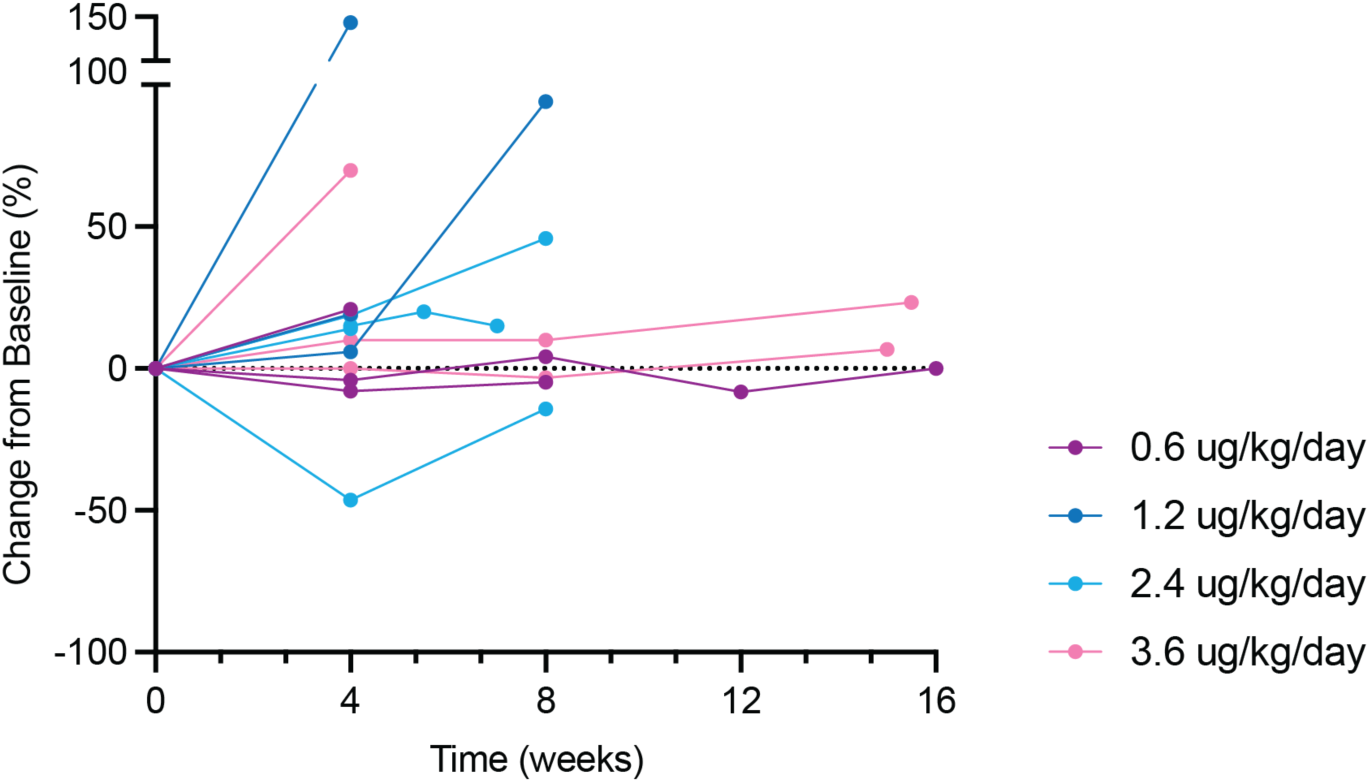
Spider plot. Percent change from baseline at time of each interval assessment (n=14) per response evaluation criteria in solid tumors (RECIST v.1.1.) calculated based on the sum of diameters for target lesions. See also Figure S1.

One patient achieved prolonged disease stability following treatment with AVB-001. The patient had primary ovarian cancer and experienced progression through three prior lines of therapy. Her disease had most recently remained stable on combination paclitaxel and bevacizumab for the preceding 10 months. However, her ability to continue was limited by side effects, including severe oral neuropathy, weight loss, and cytopenia. Because of this, she enrolled in the trial and received AVB-001 at dose level 4. Following drug administration, she subsequently remained off treatment for six months (182 days), during which time all side effects from prior chemotherapy resolved. Follow-up scans through month 4 confirmed ongoing disease stability with progression ultimately noted at the end of month 5 (169 days post-treatment with AVB-001).

### Observed Pharmacokinetics

The level of IL-2 was observed via serial plasma concentration assessment at baseline, and at 12-, 24-, 72-, 168-, 336-, 504-, and 648-hours following AVB-001 administration. Plasma IL-2 levels peaked at 24-hours (1 day) post-administration, which was followed by a steady decline in concentration and a return to baseline levels by 336-hours (7 days) post-administration (**Figure 4A**). Peak serum IL-2 concentrations increased linearly with respect to dose (R2 = 0.7413) (**Supplemental Figure 2**). To estimate the levels of IL-2 in the peritoneal cavity following AVB-001, a two-compartment mathematical model was developed, and individual patient parameters were estimated with nonlinear effects modeling (**Supplementary Figures 3 & 4, Supplementary Tables 1-4, Supplementary Text**). Model fits were in good agreement with measured serum IL-2 concentrations (**Figure 4C-F**), and modeled predictions of IL-2 in the peritoneal cavity were comparable to the amounts administered in a Phase I clinical trial of peritoneal infusions of recombinant IL-2 (**Figure 4G-J**) ^22^. Intraperitoneal concentrations could not be predicted with high confidence due to the uncertainty of peritoneal fluid volumes. However, concentrations estimated based on varying peritoneal volumes reveal that even in cases of significant intraperitoneal fluid accumulation, IL-2 concentrations are at least 7x higher in the peritoneal cavity compared to the blood (**Supplementary Table S5**). Furthermore, our results illustrate that patient-to-patient differences in capsule longevity best explain inter-patient variability (**Supplementary Figure 5**).

**Fig. 4.**
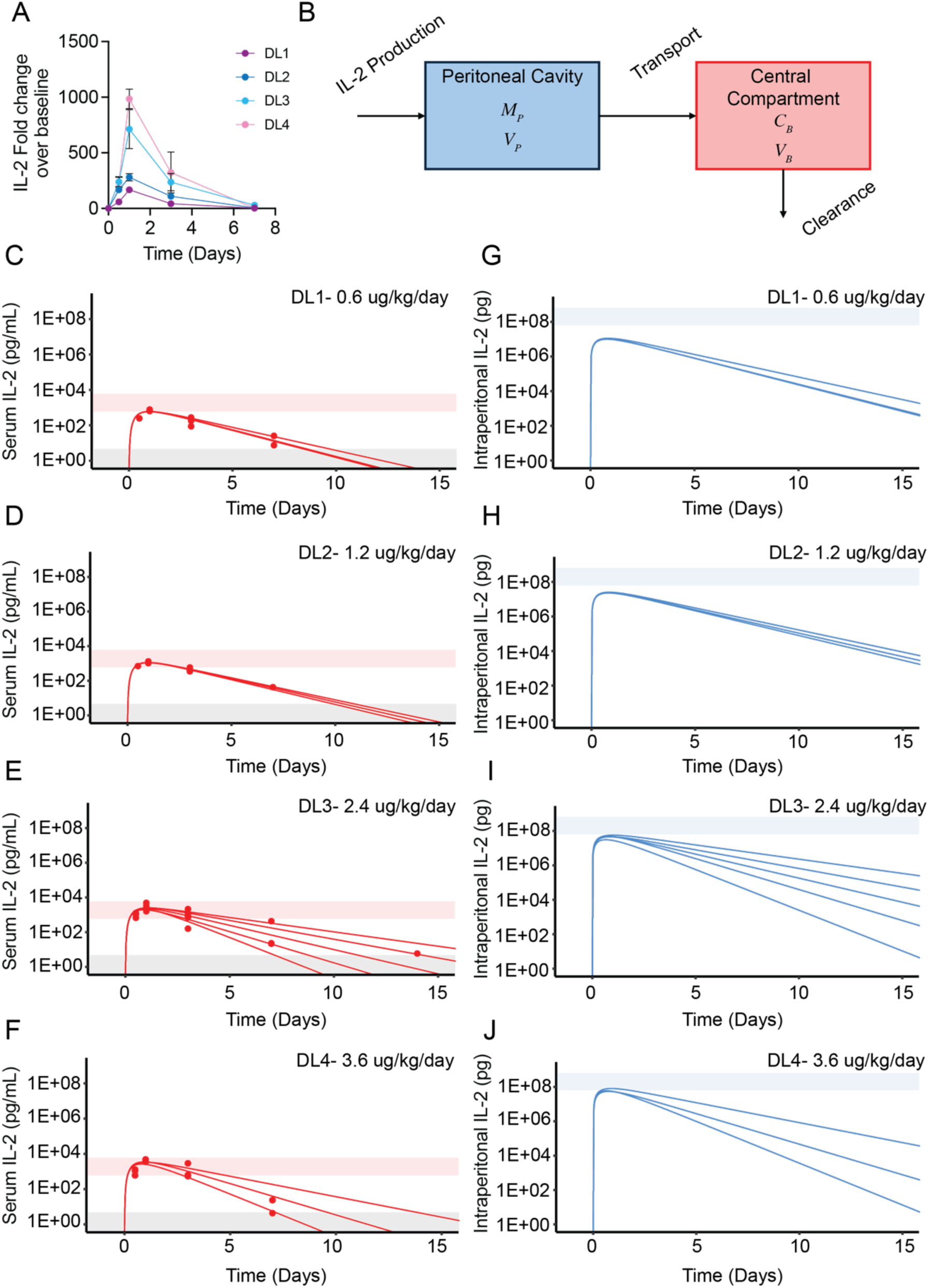
Serum IL-2 pharmacokinetics and intraperitoneal modeling. (A) Plasma concentration of IL-2 measured longitudinally. Data are expressed as mean ± SEM. (B) Schematic of the two-compartment model of AVB-001 treatment. *M*_*p*_: mass of IL-2 in the peritoneal cavity (pg). *C*_*B*_: Concentration of IL-2 in central compartment (pg/mL). (C-F) Model fits of serum IL-2 levels overlaid with individual patient data at each dose level. Grey shading represents the assay’s lower limit of quantification. The red shading denotes serum IL-2 concentrations reported by Edwards et al. for 7-day infusions of 6E5 IU/m²/day and 6E6 IU/m²/day. (G-J) Modeled predictions of intraperitoneal IL-2 levels over time for each dose level. Blue shading represents the absolute IL-2 doses administered in Edwards et al., corresponding to 7-day infusions of 6E5 IU/m²/day and 6E6 IU/m²/day (See also Figures S2-S5, Tables S1-S5, and Supplementary text).

### Clinical Evidence of Immunological Pharmacodynamics

To investigate the immunogenicity of AVB-001, peripheral blood from each patient was evaluated for changes in immune cell frequencies, as well as fluctuations in protein biomarkers. Notably, flow cytometric analysis of peripheral immune cells demonstrated an increase in circulating CD8+Ki67+, CD4+Ki67+, and NK+Ki67+ effector cells (**Figure 5A-D**). Across each cell population, the percent of cells peaked at day 8 post-transplant and returned to baseline levels by day 14. Changes in frequency were not dose-dependent. Importantly, there was no notable increase in the percentage of CD4+ T-regulatory cells, which is a common disadvantage of systemic IL-2 therapy.

**Fig. 5.**
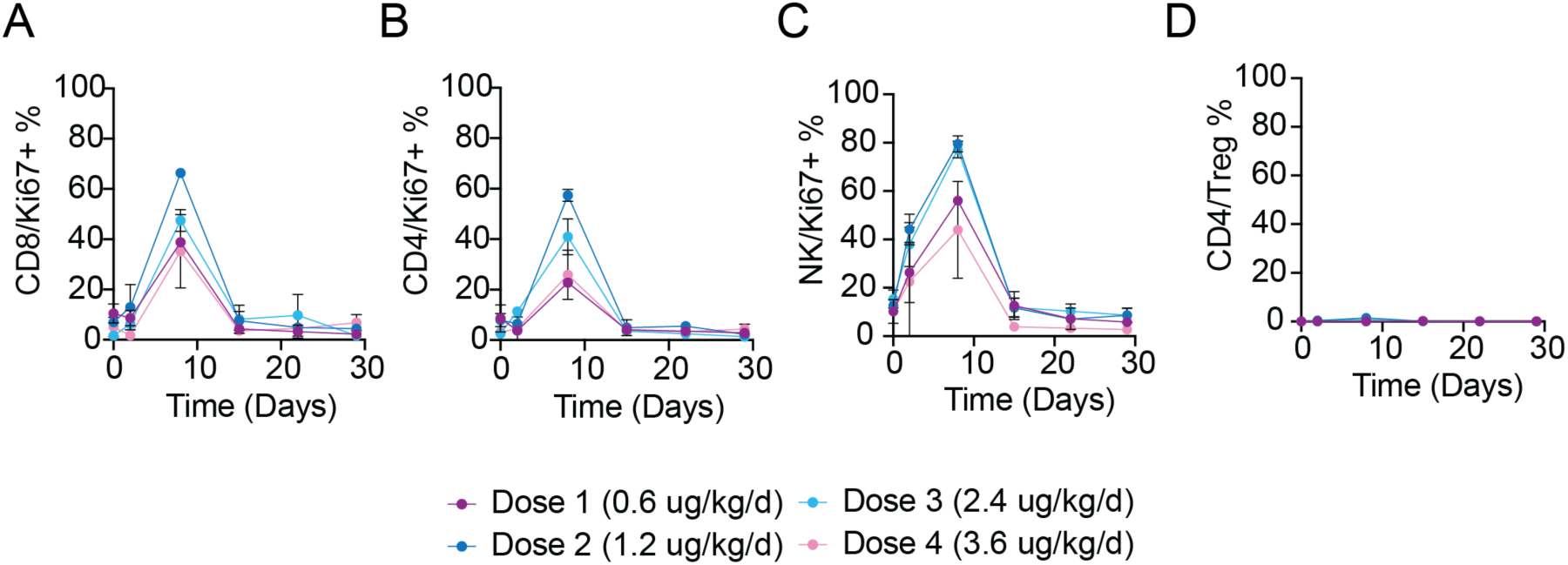
Peripheral blood immune cell analysis. Flow cytometric analysis of **(A)** CD8⁺Ki67⁺ and **(B)** CD4⁺Ki67⁺ proliferating T cells, **(C)** CD56⁺Ki67⁺ NK cells, and **(D)** CD4⁺FOXP3⁺ T regulatory cells in peripheral blood at days 0, 2, 8, 15, 22, and 29 following capsule transplantation. Data are expressed as mean ± SEM. See also Table S6 and S7.

A well-documented phenomenon following immunotherapy is the upregulation of surface immune checkpoint receptors (e.g., PD-1, CTLA-4, TIM-3, LAG-3, TIGIT) as a homeostatic mechanism to regulate immune activation and prevent excessive inflammation or autoimmunity^25^. Further analysis of effector immune populations revealed a dose-dependent trend toward increased PD-1 and TIM-3 expression, particularly when considering maximum fold change. However, statistical significance was only observed for CTLA-4 expression (**Figure 6A-C**). While co-expression of multiple checkpoint receptors is often associated with T-cell exhaustion, the isolated upregulation of CTLA-4 has been reported as an early activation marker rather than a sign of functional decline^26,27,28^.

**Fig. 6.**
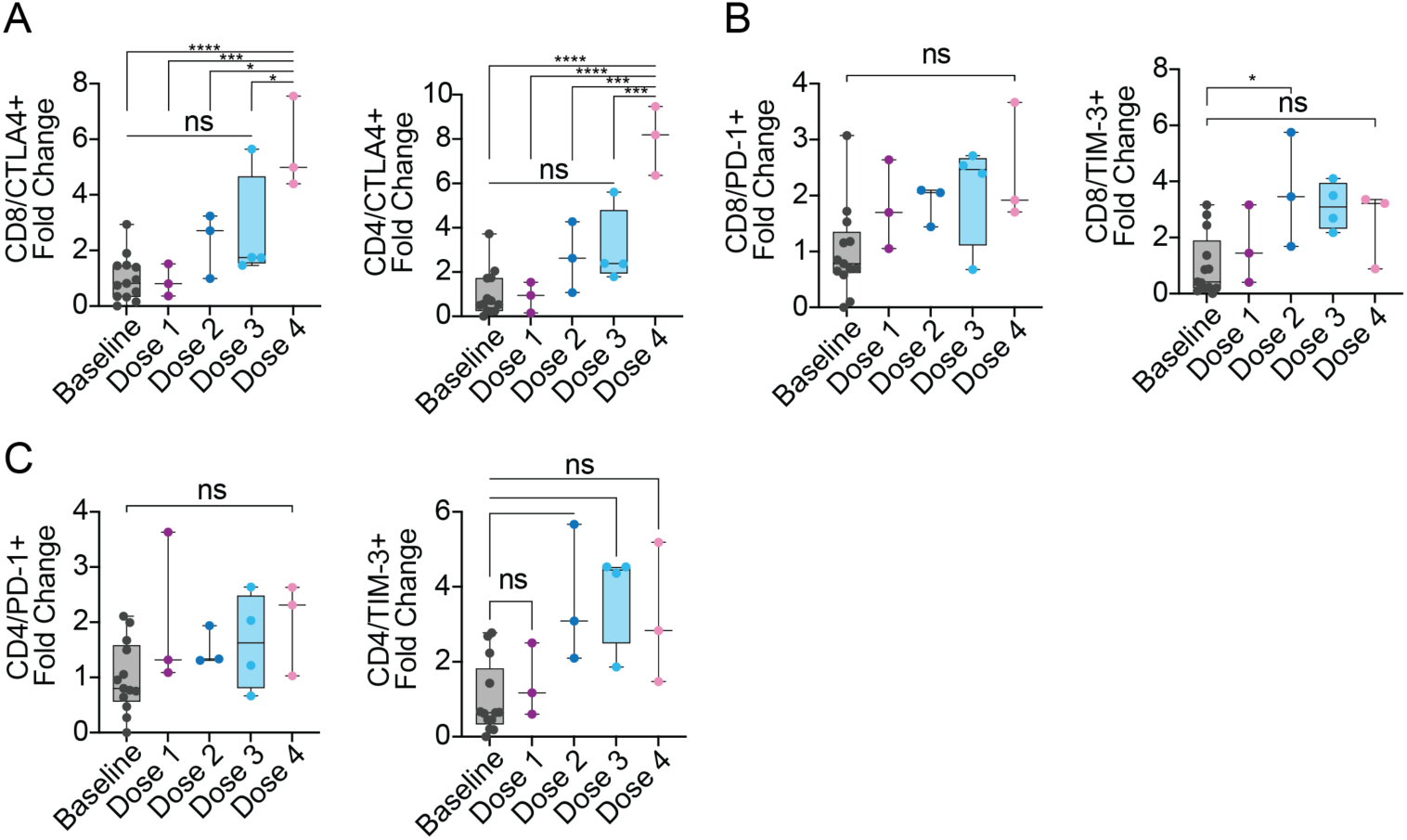
Cell surface immune checkpoint expression. Flow cytometric analysis of peak expression of surface immune checkpoint molecules **(A)** CTLA-4 **(B)** PD-1 and **(C)** TIM-3 on peripheral blood CD8⁺ and CD4⁺ T cells compared to baseline. Data are expressed as mean ± SEM. See also Table S6 and S7.

Evaluation of serum biomarkers demonstrated robust immune activation, with dose-dependent increases observed in IFN-γ, IP-10, CD25, and IL-6 (**Figure 7A-D**). Peak elevations of IFN-γ and IL-6 occurred at day 2 post-treatment, whereas IP-10 and CD25 peaked at day 8. While IFN-γ, IP-10, and IL-6 levels returned to baseline, CD25 remained elevated until day 30, particularly in dose levels 3 and 4, suggesting sustained immune activation at higher doses. TNF-α levels also increased; however, the kinetics varied across dose cohorts, with dose levels 1 and 3 peaking at day 2, dose level 2 peaking at day 8, and dose level 4 peaking at day 5 (**Figure 7E**). Finally, CA-125, a clinically relevant biomarker for ovarian cancer burden, exhibited an inverse correlation with AVB-001 dose. Higher AVB-001 doses were associated with attenuated increases in CA-125 by day 29, though this trend did not reach statistical significance (**Figure 7F**).

**Fig. 7.**
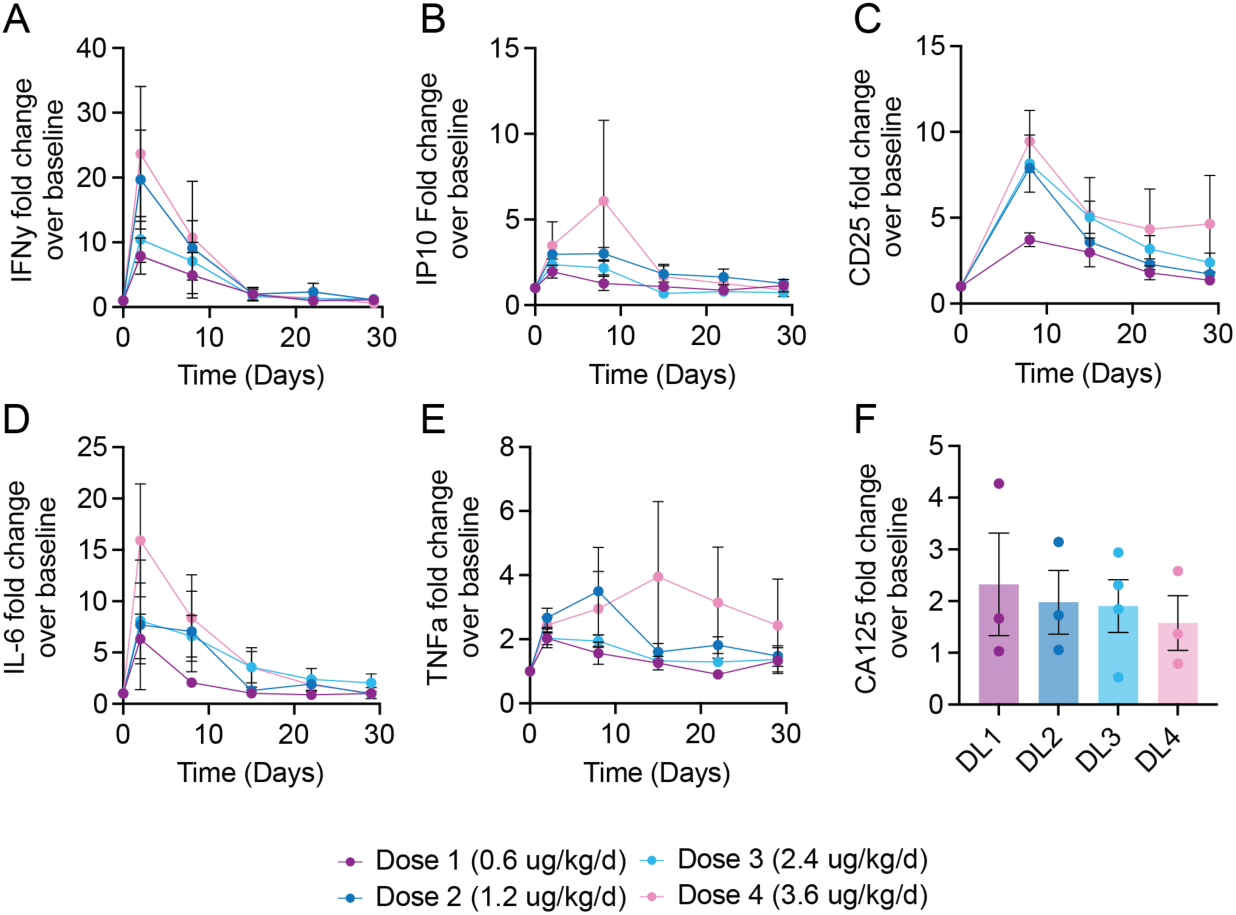
Analysis of Plasma Biomarkers Post-Capsule Transplantation. Fold change over baseline of plasma concentration of **(A)** IFN-γ, **(B)** IP-10, **(C)** CD25, **(D)** IL-6, **(E)** TNF-α, measured longitudinally, and (F) CA125 on day 29. Data are expressed as mean ± SEM. See also Figure S6.

Complete blood cell counts were evaluated for elevated eosinophils, platelets, and lymphocytes as secondary metrics for inflammatory response, coagulation activity, and immune activation, respectively. Increases were observed across all three cell types, though only eosinophils showed a marked elevation above the normal range, which is an expected effect of IL-2 treatment (**Supplementary Figure 6A-C**). This rise is consistent with IL-2’s role in promoting eosinophil survival and expansion, likely driven by secondary cytokine signaling. Liver function was monitored via serum levels of AST, ALT, and GGT over a 30-day period. Across all dose groups, transient increases were observed shortly after dosing. However, these elevations returned to baseline by Day 15–20, and no sustained or dose-dependent hepatotoxicity was evident. These findings indicate that treatment was generally well tolerated from a hepatic standpoint, with only transient and reversible perturbations in liver enzymes at higher doses (**Supplementary Figure 6D**).

These results, while demonstrating robust pharmacodynamic activity, indicate that the immunological response to a single dose of AVB-001 is transient, with most immune biomarkers and effector cell populations returning to baseline within two weeks. This short duration of activity, while promising, underscores the potential need for repeated dosing to sustain immune engagement. Given the minimally invasive intraperitoneal delivery method, the lack of observed anti-therapy antibody formation in preclinical murine models, and the absence of T-regulatory cell expansion seen clinically, we hypothesize that multiple doses of AVB-001 may prolong the activation of cytotoxic T cells and NK cells, thereby enhancing therapeutic efficacy. To investigate the safety and feasibility of a repeated dosing regimen, we next evaluated AVB-001 re-administration in non-human primates.

### Pharmacology and Safety Following the Second Administration of AVB-001 in Non-Human Primates

To evaluate the potential toxicity and safety of a second dose of AVB-001, non-human primates were administered 10 μg/kg/day of AVB-001 hIL-2 on day 0, followed by a second dose on day 28 via intraperitoneal delivery using a laparoscopic technique (**Figure 8A**). The study concluded 120 days after the first administration (**Figure 8B**). Blood samples were collected at multiple timepoints and physiological parameters, including weight and body temperature, were monitored to assess the overall health of the animals.

**Fig. 8.**
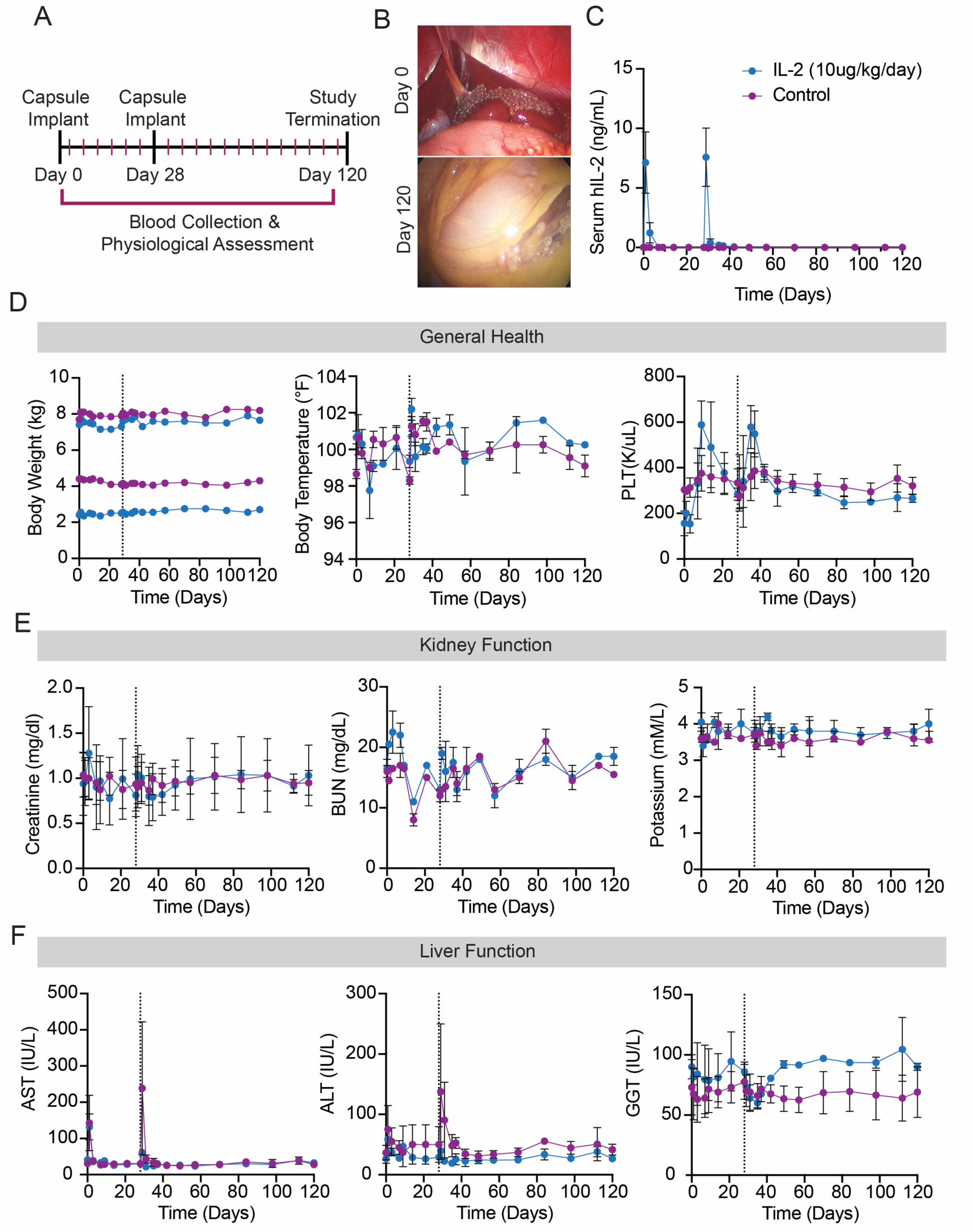
**Pharmacological assessment of AVB-001 re-administration in Cynomolgus macaques**. **(A)** Study overview timeline. **(B)** Representative images of the intraperitoneal space immediately following capsule implantation (Day 0, left) and at study termination (Day 120, right). **(C)** Serum concentrations of human IL-2 (hIL-2) over time. **(D)** General toxicity evaluation, including changes in body weight (left), temperature (middle), and platelet counts (right) over time. **(E)** Kidney function assessment via changes in creatinine levels (left), blood urea nitrogen (BUN) levels (middle), and potassium levels (right) over time. **(F)** Evaluation of liver function through changes in AST (left), ALT (middle), and GGT (right) levels over time. The dashed line indicates the timing of the second dose administration. (n = 2 per group) Data are expressed as mean ± SEM. See also Figure S7.

Serial blood collections were conducted on days 0, 1, 3, 7, 9, 14, 21, 28, 29, 31, 35, 37, 42, 49, 56, biweekly thereafter, and on the day of termination. Serum concentrations of human IL-2 (hIL-2) were analyzed at each timepoint. As expected, hIL-2 was undetectable in saline-treated animals. In AVB-001-treated animals, hIL-2 was transiently detectable in the serum following each dose, with peak levels observed on day 1 post-administration (mean IL-2 levels on Day 1: 7,134.5 ± 3,639.4 pg/mL; Day 29: 7,583.5 ± 3,463.4 pg/mL). There were no significant differences in the kinetics of hIL-2 secretion between the first and second administrations, with serum levels consistently returning to baseline within 7–14 days post-dose (**Figure 8C**).

To assess physiological response to capsule administration and re-dosing, animal body weights and temperature were evaluated over the course of the study. No significant difference was evaluated when comparing AVB-001 treated primates to saline controls indicating that the treatment was generally well tolerated (**Figure 8D).**

Hematologic assessment revealed normal results across most measurements in vehicle-treated animals with a small number of occasions where leukocyte values (e.g. % lymphocytes, % neutrophils, or total WBC) were slightly out of range for only a single timepoint. However, these changes were transient, did not correlate with any other observations or findings, and are not of clinical concern (**Figure 8D, Supplemental Figure 6)**.

Liver function tests revealed only mild, transient AST and ALT elevations—some also present in controls—likely related to surgical procedures rather than treatment. GGT values stayed within healthy ranges (**Figure 8E**). Kidney function parameters (creatinine, BUN, potassium) remained stable and within 20% of normal for the study’s duration, indicating no adverse renal effects (**Figure 8F**).

## DISCUSSION

This first-in-human phase I dose escalation trial investigates the use of encapsulated cell technology to employ sustained and self-limited intraperitoneal production of single-agent IL-2 in patients with platinum-resistant HGSOC. At the evaluated doses, a single administration of AVB-001 resulted in an ORR of 7.1% with DOR of 29 days. Fifty percent of patients experienced a period of stable disease; median duration of clinical stability was 2.57 months with one patient at the highest dose level experiencing off-treatment disease stabilization for 6 months. The pharmacokinetic results further support the clinical activity of this agent even among a heavily pretreated cohort. After IP administration of the study agent, we observed a linear dose-dependent rise in plasma IL-2 concentration when measured serially. This rise peaked after a period of 24 hours until it subsequently declined to baseline levels by day 7. In addition to the expected rise in plasma IL-2 levels after IP administration, further evidence of the clinical activity of AVB-001 is demonstrated by immune system activation that persisted for approximately two weeks. Specifically, flow cytometric analysis revealed an increase in circulating CD4+, CD8+, and NK effector cell proliferation that was not accompanied by a concomitant rise in CD4+ T-regulatory cells. These findings are important because they suggest that, unlike systemically administered IL-2, which has previously been associated with an increase in CD4+ T-regulatory cells that serve to suppress the potential immune response generated by the study agent, IP administration of AVB-001 selectively increases immune effector cells, which could perpetuate greater immune activation^21^. Further evidence of the immunogenicity of this compound is exhibited by dose-dependent rises in IFN-γ, IL-10, CD25, and IL-6. Taken together, these findings propose an immune-based mechanism that could provide meaningful clinical benefit in a subset of patients within this cohort.

As has been previously observed with the administration of IL-2 based immunotherapy, the administration of AVB-001 led to upregulation of surface immune checkpoint receptors^29^. Immune checkpoint receptors (i.e. PD-1, CTLA-4, LAG-3, TIM-3, TIGIT) play a key role in the prevention of excessive immune activation. Within our study group, this phenomenon is observed via the dose-dependent upregulation of CTLA4+ CD4 and CD8 T cells in peripheral blood. This upregulation suggests that combining AVB-001 with CTLA-4 antibody blockade may further potentiate the immunogenicity of the compound by limiting intrinsic suppressive checkpoint mechanisms. Checkpoint blockade has been widely studied across various cancer types, and at present, there are two anti-CTLA-4 antibodies available for clinical use^30,31^. Neither, however, is currently approved for use in ovarian cancer and all approvals are for either single-agent administration or combination therapy with an anti-PD-1 or anti-PD-L1 antibody. Combination anti-PD1 checkpoint inhibition and IL-2 therapy (nemvaleukin) demonstrated encouraging early results in the context of platinum-resistant ovarian cancer (PROC) as part of the phase I/II ARTISTRY-1 trial, which reported an ORR of 21% among 14 patients with PROC^32^. These findings support the idea that, if able to reach a clinically active dose level and maintain safety, combination checkpoint inhibition and IL-2 therapy could lead to meaningful clinical benefit in PROC and other tumor types that may present as secondary metastatic disease in the peritoneal cavity and benefit from both localized and systemic immune activation.

In this trial, AVB-001 was found to be safe at the dose levels explored. The MTD and RP2D were not reached. One patient experienced a bowel perforation that was initially reported as a TRAE and a DLT. However, following a thorough clinical review, it was determined to be unrelated to AVB-001. Importantly, four additional patients were subsequently dosed at the same level without recurrence of this event, and three patients received a higher dose, further supporting that bowel perforation is unlikely to be an expected adverse event associated with IL-2 delivery via AVB-001. Building on the strong safety profile demonstrated in both human and prior non-human primate (NHP) studies, the next phase of clinical development will focus on reinitiating the phase I trial at an increased IL-2 dose^13^. This escalation aims to refine our understanding of AVB-001’s therapeutic window by identifying the MTD, any DLTs, and the RP2D. Additionally, an expansion of the phase 1 trial will incorporate repeated administration at the RP2D to better model long-term IL-2 delivery and align more closely with the 16-week regimen used in Edwards et al^22^. However, unlike the prolonged, continuous IL-2 infusions used in that study, AVB-001 represents a significantly improved delivery approach due to its novel constitutive IL-2 production capabilities.

Intraperitoneal modeling data suggest that the highest dose of AVB-001 results in local IL-2 levels comparable to the maximum tolerated dose in the seven-day infusion regimen evaluated by Edwards et al. ^22^. Importantly, no dose-limiting toxicities were observed at this dose level, despite serum IL-2 levels reaching concentrations previously associated with toxicity in the Edwards trial. Furthermore, based on pre-clinical studies in NHPs where doses up to 3x higher than the highest dose tested in this clinical study were administered, we anticipate that higher doses of AVB-001 will maintain a favorable safety profile. This would allow for the administration of IL-2 at doses higher than previously explored while also permitting re-dosing and combination with other targeted immunotherapy agents such as checkpoint inhibitors—something not feasible with prior systemic and local IL-2 regimens due to toxicity limitations^33,34^. This approach is further supported by our NHP data demonstrating stable pharmacokinetics and tolerability across multiple doses, providing a robust foundation for assessing whether extended exposure enhances immune activation and tumor control without increasing toxicity. Future trials will also include systematic collection of intraperitoneal fluid and tumor biopsies, enabling a detailed evaluation of local tumor and peritoneal immune dynamics through immune cell characterization and immune signaling biomarker collection folowing AVB-001 administration. These insights will be critical for optimizing dosing strategies and informing potential combination therapies in subsequent trial phases.

To enhance the clinical scalability of AVB-001, future efforts will also focus on optimizing the manufacturing process related to this allogeneic cell based product. Increasing the per cell production rate of hIL-2 will reduce the number of cells and thus volume of capsules per dose required to reach a target level of IL-2 and enable cell expansion of multiple doses in parallel. Additionally, integrating high-throughput automated encapsulation to improve batch consistency will decrease the need for a technician to control voltage and enable efficient high throughput production of drug product at scale. These advancements will enable broad clinical availability, reduce production bottlenecks, and dramatically reduce cost of goods per dose, ultimately supporting the transition of AVB-001 from early-phase trials to larger, multicenter studies and ultimately commercialization. Together, these advancements will refine AVB-001’s clinical potential and accelerate its path toward broader therapeutic application.

In conclusion, this study demonstrates that intraperitoneal administration of AVB-001 is safe, feasible, and shows potential for meaningful clinical activity in a cohort of patients with platinum-resistant HGSOC. We found that peritoneal delivery of AVB-001 triggered a systemic immunogenic response as evidenced by a rise in peripheral immune effector cells and inflammatory cytokines following administration. We further observed a dose-dependent upregulation of the CTLA-4 checkpoint on circulating CD8+ and CD4+ T cells, suggesting a role for combination therapies involving cytokines as priming agents to engage more effective checkpoint inhibition.

Key Resources Table.

**Key Resources Table**
| REAGENT or RESOURCE | SOURCE | IDENTIFIER |
| --- | --- | --- |
| Coaxial Needle, 18/22 Gauge | SKE | EFA035 |
| **Chemicals** |  |  |
| SLG20 Alginate | Novamatrix | 4202001 |
| Barium Chloride | Sigma-Aldrich | 342920 |
| D-Mannitol | Sigma-Aldrich | 63560 |
| 0.9% Saline, USP | Cytiva | Z1376 |
| Dulbecco's Phosphate Buffered Saline, Modified, without calcium chloride and magnesium chloride | Sigma-Aldrich | D8537 |
| HEPES (N-2-hydroxyethylpiperazine-N-2-ethane sulfonic acid) 1M solution | ThermoFisher | 15630106 |
| Potassium Chloride | Sigma-Aldrich | 60128 |
| Potassium Phosphate Monobasic | Sigma-Aldrich | 60218 |
| Magnesium Sulfate Heptahydrate | Sigma-Aldrich | 63138 |
| Magnesium Chloride Hexahydrate | Sigma-Aldrich | 63068 |
| Sodium Chloride | Sigma-Aldrich | S7653 |
| Tryple Select | ThermoFisher |  |
| Tween 20 | Sigma-Aldrich | P2287 |
| DMEM/F-12, HEPES, no phenol red | ThermoFisher | 11039047 |
| Critical commercial assays |  |  |
| Human IL-2 ELISA Kit - Quantikine | R&D Systems | D2050 |
| BSA ELISA Kit | Cygnus Technologies | F030 |
| Via1-Cassette | Chemometec | 941-0012 |
| Clinitrol 290 Reference Solution | Advanced Instruments |  |
| LAL Cartridge 5-0.05 | Charles River Laboratories | PTS2005F |
| LAL Cartridge 1 – 0.01 | Charles River Laboratories | PTS2001F |
| LAL Cartridge 0.5 – 0.005 | Charles River Laboratories | PTS20005F |
| ACT/ALERT i NST (Anaerobic) | Bomerieux | 10298 |
| BACT/ALERT i AST (Aerobic) | Bomerieux | 10297 |
| **Experimental models: Cell lines** |  |  |
| Retinal Pigment Epithelial Cells | ATCC | CRL2302 |
| **Experimental models: Organisms/strains** |  |  |
| Macaques | DSP and Worldwide Primates |  |
| **Recombinant DNA** |  |  |
| IL-2 Plasmid | Aldevron | Human IL-2 |
| **Software and algorithms** |  |  |
| R V4.4.2 | NA | NA |
| nlmxir2 V3.0.2 | NA | NA |
| Graphpad Prism V10.2.2 | NA | NA |

## Author contributions

**Conceptualization:** A.A.J., S.W., O.V., C.D.U.

**Formal Analysis:** C.H., S.A.F., H.D.C., J.D.

**Funding Acquisition**: O.V.

**Investigation:** J.S., J.C.A.M., C.H., A.N., O.Y., A.M.B., C.M., A.A.J., S.N.W., T.S., K.R., K.A., L.J., J.O., P.R., M.J., T.T.S., I.J., D.L.

**Software:** J.D., O.A.I.

**Methodology:** C.D.U., C.H., R.N., O.V., A.A.J.

**Supervision:** C.D.U, O.V., R.C., A.A.J., O.A.I., S.W.

**Visualization:** S.A.F., H.D.C., J.D.

**Writing – Original Draft:** S.A.F., H.D.C., J.D., J.S.

**Writing – Review & Editing:** S.A.F., H.D.C., J.D., J.S., O.A.I., O.V., R.C., A.A.J., S.W., C.D.U.

## Acknowledgements

The study was funded by the following: NIH MD Anderson Cancer Center Support Grant (#P30CA016672; Institutional Tissue Bank and ORION), NIH MD Anderson T32 Training Grant (#T32CA101642), Cancer Prevention Research Institute of Texas (RR160047), Advanced Research Projects Agency for Health (ARPA-H) (AY1AX000003), Common Fund (AI177915), and Sponsored Research Agreement Avenge Bio (#AVB-001).

This study was supported in part of the Translational Molecular Pathology-Immunoprofiling Lab (TMP-IL) Platform at the Department Translational Molecular Pathology, the University of Texas MD Anderson Cancer Center.

This research was supported [in part] by the Intramural Research Program of the National Institutes of Health (NIH). The contributions of the NIH author(s) were made as part of their official duties as NIH federal employees, are in compliance with agency policy requirements, and are considered Works of the United States Government. However, the findings and conclusions presented in this paper are those of the author(s) and do not necessarily reflect the views of the NIH or the U.S. Department of Health and Human Services.

