## Supplemental Materials for "First-In-Human Trial of Encapsulated Cell-Based Protein Producers for Localized IL-2 in Patients with High-Grade Serous Ovarian Carcinoma"

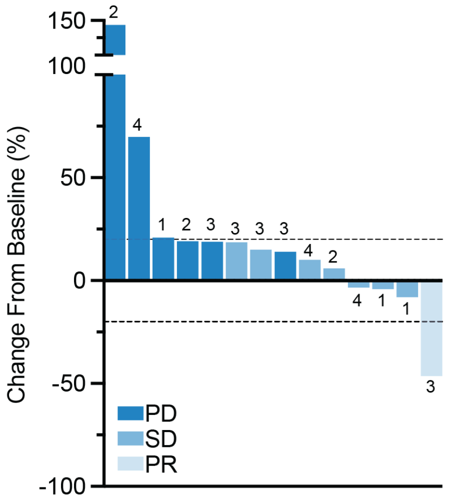

**Supplemental Fig. 1. Waterfall plot. Related to Figure 3**

Best response change from baseline (% change, n=14) per response evaluation criteria in solid tumors (RECIST v.1.1.) by cohort, calculated based on sum of diameters for target lesions. Value above each bar represents dose cohort. (1- 0.6 ug/kg/day; 2- 1.2 ug/kg/day; 3- 2.4 ug/kg/day; 4- 3.6 ug/kg/day).

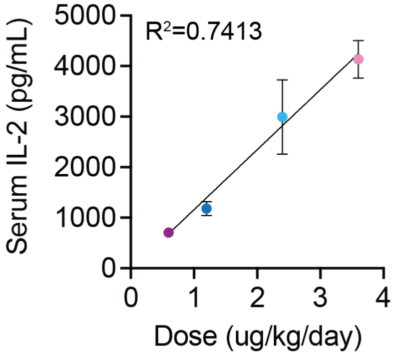

**Supplementary Fig. 2. IL-2 linearity, related to Figure 4.** Each dose of AVB-001 administered resulted in a proportional, linear increase in serum IL-2 levels. Data are presented as mean ± SEM**.**

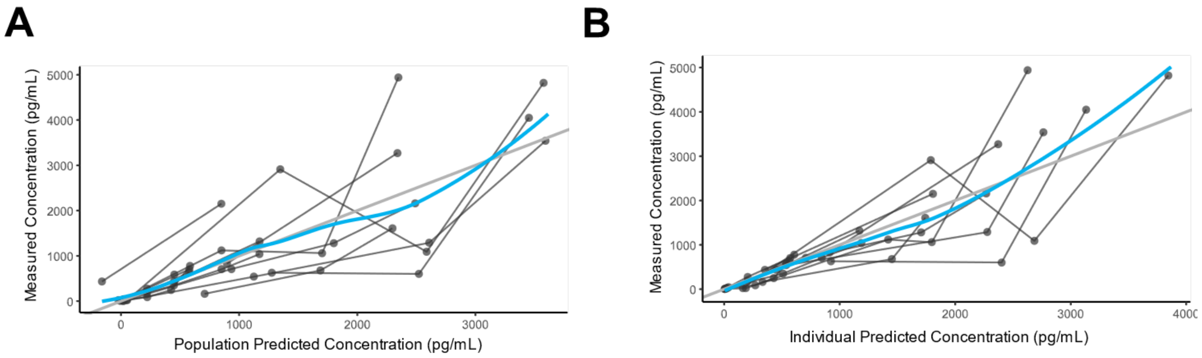

**Supplementary Fig.3. Assessment of model performance with all parameters as random effect variables, related to Figure 4.** A) Comparison of experimental data to population model predictions. B) Comparison of experimental data to individual model predictions. Blue curves: linear regression of individual points.

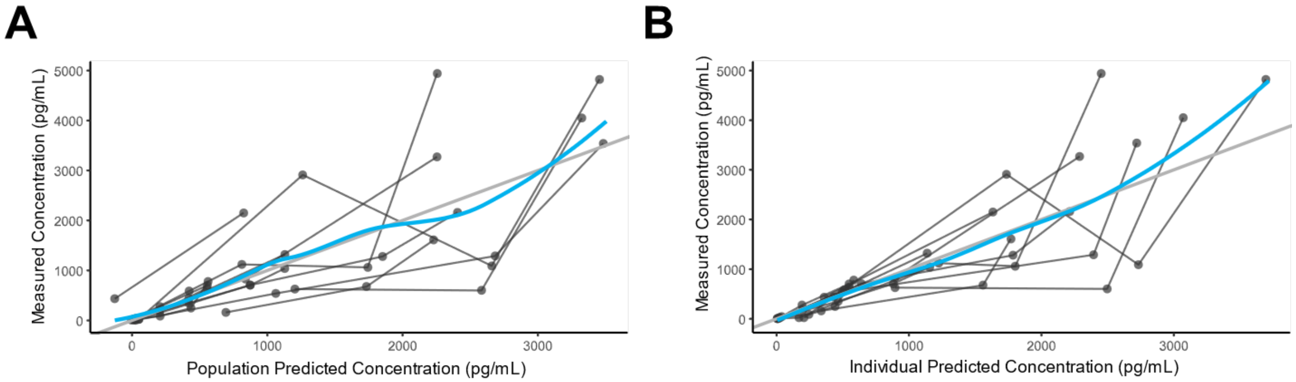

**Supplementary Fig. 4. Assessment of model performance with only $\lambda$ as a random effect variable, related to Figure 4.** A) Comparison of experimental data to population model predictions. B) Comparison of experimental data to individual model predictions. Blue curves: linear regression of individual points.

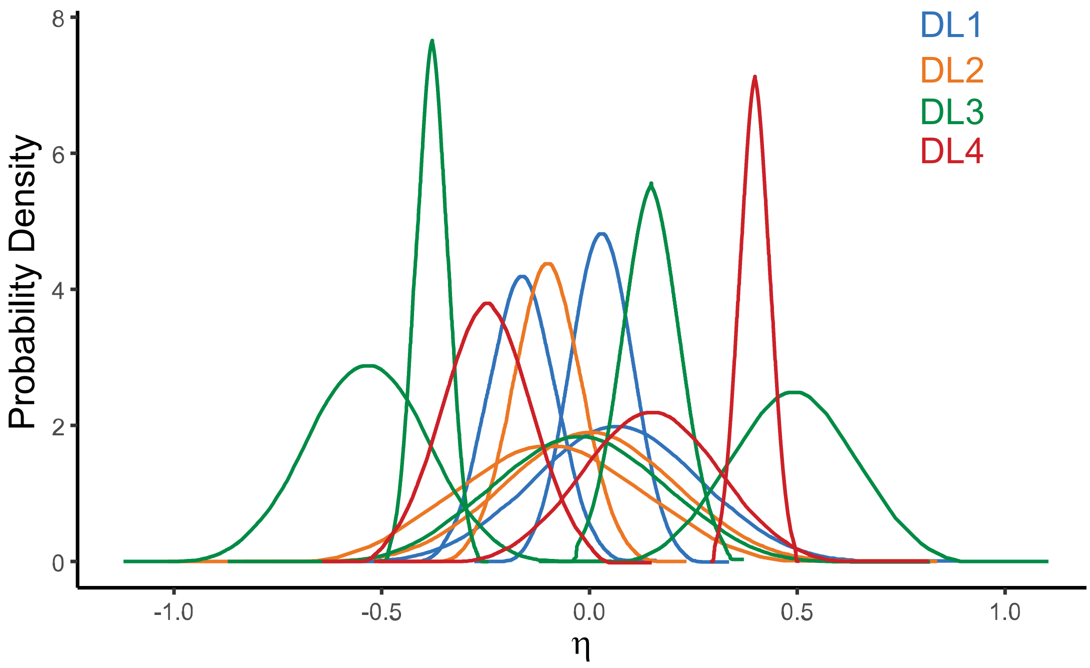

**Supplementary Fig. 5. Empirical Bayes estimates of individual λ values in final model, , related to Figure 4.** X axis is given in terms of difference from population value in log space (η). Colors represent different dose levels (DL).

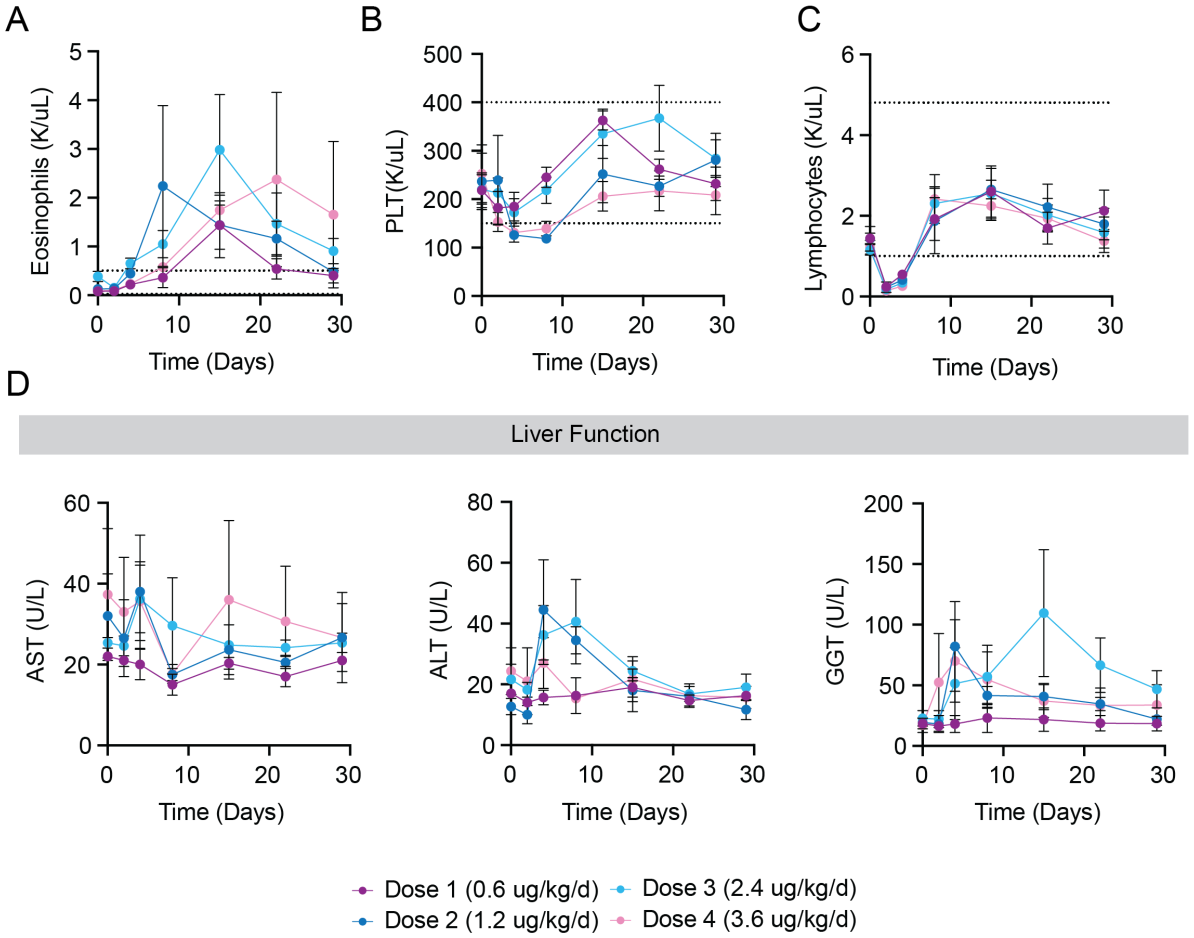

**Supplementary Fig. 6. Longitudinal analysis of CBC and liver function post treatment, related to Figure 7. (A)** Total eosinophil **(B)** platelet and **(C)** lymphocyte counts quantified at days 0, 2, 4, 8, 15, 22, and 29 post-treatment (n = 3–5 per group). **(D)** Liver function assessed through the analysis of AST, ALT, and GGT levels over time at the same intervals (n = 3–5 per group). Data are expressed as mean ± SEM.

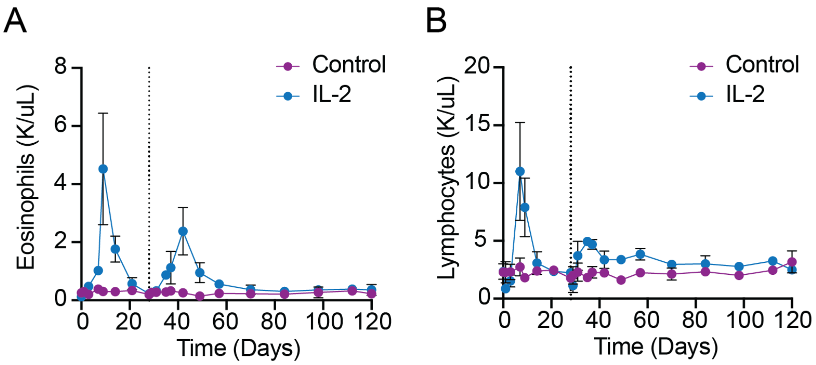

**Supplemental Fig. 7. Long-Term Tracking of Eosinophil and Lymphocyte Counts Post-Treatment, related to Figure 8. (A)** Total eosinophil and **(B)** lymphocyte count quantified serially for the duration of study (Day 0 - 120) post-treatment (n = 2 per group). Data are expressed as mean ± SEM.

**Supplementary Table S1. Related to Figure 4.** Description of model parameters.

| Parameter | Meaning | Source |
|---|---|---|
| $N_0$ | Initial number of capsules administered. Set as initial condition for model simulation.<br>Units: # | Set by individual patient data. |
| $k_{prod}$ | IL-2 production rate per capsule.<br>Units: pg/day | Set by individual patient data. |
| $BW$ | Patient body weight.<br>Units: kg | Set by individual patient data. |
| $BW_{median}$ | Median body weight of all patients. Fixed as 66.6 kg.<br>Units: kg | Set by all combined patient data. |
| $\lambda$ | First-order capsule decay rate constant.<br>Units: days$^{-1}$ | Estimated from data. |
| $k_{BP}$ | First order IL-2 peritoneal transport rate constant.<br>Units: days$^{-1}$ | Estimated from data. |
| $k_{clear,0}$ | Population value for IL-2 systemic clearance rate constant, scaled by body weight. | Estimated from data. |

| | |
|---|---|
| | Units: $days^{-1}$ |
| $V_{B,0}$ | Population data for volume of central compartment, scaled by body weight.<br>Units: mL |

**Supplementary Table S2. Related to Figure 4.** Estimated model parameters with all parameters set as random effects. Values are reported to two significant digits.

| Parameter | Population Estimate | Between Subject Variability | Shrinkage |
|---|---|---|---|
| $\lambda$ | $0.73\ days^{-1}$ | 34 % | 11% |
| $k_{BP}$ | $1.8\ days^{-1}$ | 4.1% | 93% |
| $k_{clear,0}$ | $6.5\ days^{-1}$ | 2.0% | 94% |
| $V_{B,0}$ | $5{,}500\ mL$ | 7.3% | 78% |

**Supplementary Table S3. Related to Figure 4.** Estimated model parameters with only $\lambda$ as a random effect variable. Values are reported to two significant digits.

| Parameter | Population Estimate | Between Subject Variability | Shrinkage |
|---|---|---|---|
| $\lambda$ | $0.67\ days^{-1}$ | 34% | 12% |
| $k_{BP}$ | $2.1\ days^{-1}$ | $n/a$ | $n/a$ |
| $k_{clear,0}$ | $7.4\ days^{-1}$ | $n/a$ | $n/a$ |
| $V_{B,0}$ | $5{,}500\ days^{-1}$ | $n/a$ | $n/a$ |

**Supplementary Table S4. Related to Figure 4.** Individual expected $\lambda$ values and corresponding treatment half-lives.

| Patient ID | $\lambda\ (days^{-1})$ | $t_{1/2}(days)$ |
|---|---|---|
| 001-001 | 0.60 | 1.2 |
| 001-003 | 0.70 | 1.0 |
| 001-004 | 0.71 | 0.99 |
| 001-006 | 0.66 | 1.1 |
| 004-002 | 0.59 | 1.2 |
| 004-003 | 0.62 | 1.1 |
| 001-007 | 0.64 | 1.1 |
| 001-008 | 0.38 | 1.8 |
| 004-005 | 1.1 | 0.64 |

| | | |
|---|---|---|
| 006-001 | 0.50 | 1.4 |
| 006-002 | 0.83 | 0.84 |
| 003-001 | 1.1 | 0.62 |
| 004-006 | 0.54 | 1.3 |
| 006-003 | 0.82 | 0.85 |

**Supplementary Table S5. Related to Figure 4.** Model predictions of maximum serum IL-2 concentration, maximum peritoneal IL-2 concentration, and mean peritoneal:serum IL-2 concentration ratio for each dose level. Values given for three peritoneal volume estimates. Values are reported to three significant digits.

| Dose Level | Peritoneal Volume (mL) | Peak Serum IL-2 Concentration (pg/mL) | Peak Peritoneal IL-2 Concentration (pg/mL) | Mean Peritoneal: Serum Ratio |
|---|---|---|---|---|
| DL1 | 50 | 595 (6.58) | 208,000 (11,800) | 350 |
| | 250 | 595 (6.58) | 41,700 (2,360) | 70.1 |
| | 2500 | 595 (6.58) | 4,170 (236) | 7.01 |
| DL2 | 50 | 1120 (16.1) | 475,000 (21,800) | 423 |
| | 250 | 1120 (16.1) | 94,900 (4,370) | 84.6 |
| | 2500 | 1120 (16.1) | 9,490 (437) | 8.46 |
| DL3 | 50 | 2280 (218) | 922,000 (174,000) | 404 |
| | 250 | 2280 (218) | 184,000 (34,800) | 80.7 |
| | 2500 | 2280 (218) | 18,400 (3,480) | 8.07 |
| DL4 | 50 | 3140 (357) | 1,300,000 (227,000) | 415 |
| | 250 | 3140 (357) | 261,000 (45,500) | 83.1 |
| | 2500 | 3140 (357) | 26,100 (4,550) | 8.31 |

**Supplementary Table S6. Related to Figures 5 and 6. Myeloid panel design**

| Surface Antibody | Fortessa X20 Channel | Laser | Company | Catalog | Clone |
|---|---|---|---|---|---|
| CD3 FITC | FITC | BLUE | Biolegend | 300406 | UCHT1 |
| CD19 FITC | FITC | BLUE | Biolegend | 302206 | HIB19 |
| CD20 FITC | FITC | BLUE | Biolegend | 302304 | 2H7 |
| CD56 FITC | FITC | BLUE | Biolegend | 318306 | HCD56 |
| CD123 PerCP-Cy5.5 | PerCP-Cy5.5 | BLUE | Biolegend | 306016 | 6H6 |
| CD14 BV421 | BV421 | VIOLET | BD Biosciences | 563743 | MΦP9 |
| Live/Dead Yellow | BV510 | VIOLET | Life Technologies | L-34968 | - |
| CD11c BV605 | BV605 | VIOLET | BD Biosciences | 563929 | B-ly6 |
| CD327 (SIGLEC6) BV650 | BV650 | VIOLET | BD Biosciences | 747911 | 767329 |
| CD16 BV711 | BV711 | VIOLET | Biolegend | 331536 | L161 |

| | | | | | |
|---|---|---|---|---|---|
| PD-1 (CD274) BV786 | BV786 | VIOLET | Biolegend | 329736 | 29E.2A3 |
| CD45 BUV395 | BUV395 | UV | BD Biosciences | 563792 | HI30 |
| CD16 BUV496 | BUV496 | UV | BD Biosciences | 612944 | 3G8 |
| PD-L2 (CD273) BUV661 | BUV661 | UV | BD Biosciences | 564924 | MIH18 |
| CD86 APC-R700 | Alexa Fluor 700 | RED | BD Biosciences | 565149 | 2331 (FUN-1) |
| CD11b APC/Fire 750 | APC-Cy7 | RED | Biolegend | 101262 | ICRF44 |
| CXCR2 (CD182) | APC | RED | BD Biosciences | 551127 | 6C6 |
| CD141 PE | PE | RED | BD Biosciences | 559781 | 1A4 |
| CD33 PE-CF594 | PE-CF594 | YG | BD Biosciences | 562492 | WM53 |
| HLA-DR PE-Cy7 | PE-Cy7 | YG | BD Biosciences | 335795 | L243 |

**Supplementary Table S7. Related to Figures 5 and 6. Function and Memory T cell panel design**

| **Surface Antibody** | **Fortessa X20 Channel** | **Laser** | **Company** | **Catalog** |
|---|---|---|---|---|
| CCR7 PerCP-Cy5.5 | PerCP-Cy5.5 | BLUE | Biolegend | 353220 |
| CD45RA V450 | BV421 | VIOLET | BD Biosciences | 560362 |
| Live/Dead Yellow | BV510 | VIOLET | ThermoFisher | L-34968 |
| ICOS SB600 | BV605 | VIOLET | ThermoFisher | 63-9948-42 |
| PD-1 (CD279) BV650 | BV650 | VIOLET | BD Biosciences | 564104 |
| Tim3 BV711 | BV711 | VIOLET | Biolegend | 345024 |
| CTLA-4 (CD152) BV786 | BV786 | VIOLET | BD Biosciences | 563931 |
| CD3 BUV737 | BUV737 | UV | BD Biosciences | 612752 |
| CD45 BUV395 | BUV395 | UV | BD Biosciences | 563792 |
| CD4 BUV496 | BUV496 | UV | BD Biosciences | 612936 |
| CD56 BUV661 | BUV661 | UV | BD Biosciences | 750478 |
| CD8 Alexa Fluor 700 | Alexa Fluor 700 | RED | BD Biosciences | 557945 |

| | | | | |
|---|---|---|---|---|
| CD25 APC-e780 | APC-e780 | RED | ThermoFisher | 47-0259-42 |
| CD27 PE | PE | YG | Biolegend | 356406 |
| CD28 PE-Cy7 | PE-Cy7 | YG | BD Biosciences | 560684 |
| **Intracellular Antibody** | **Fortessa X20 Channel** | **Laser** | **Company** | **Catalog** |
| FoxP3 PE-CF594 | PE-CF594 | YG | Biolegend | 320126 |
| TCF7/TCF1 Alexa Fluor 488 | FITC | BLUE | BD Biosciences | 567018 |
| Ki67 APC | APC | RED | ThermoFisher | 17-5699-42 |

**Supplementary Table S8. Representativeness of study participants**

| Cancer type/subtype/stage/condition | Platinum-resistant high-grade serous ovarian, fallopian tube, or peritoneal adenocarcinoma |
|---|---|
| Considerations related to: | |
| Sex | This disease affects the organs of the female reproductive tract and thus occurs exclusively in females. |
| Age | The median age at diagnosis for ovarian cancer is 63 years old. The highest rates are observed in women between the ages of 55 and 64. |
| Race/ethnicity | The overall incidence of ovarian cancer is 10.3 cases per 100,000 persons. The highest incidence is seen in Non-Hispanic American Indian/Alaskan Native women at 11.4 cases per 100,000 persons and the lowest incidence is seen in Non-Hispanic Black women at 9.2 cases per 100,000 persons.<br><br>In terms of death rate, the overall death rate is 5.9 deaths per 100,000 persons. The highest death rate is seen in Non-Hispanic American Indian/Alaskan Native women at 6.3 cases per 100,000 persons and the lowest death rate is seen in the Non-Hispanic Asian/Pacific Islander population. |
| Geography | In the United States, approximately 20,890 cases of ovarian cancer will be diagnosed and 12,730 patients will die in 2025. |
| Overall representativeness of this study | The small sample size of this population limits generalizability to the broader population. |

| | |
|---|---|
| | The age distribution skews slightly older than the literature, with a median age at enrollment of 68 years.<br><br>The racial/ethnic composition of this study is not reflective of the national population, as this study includes only women of White race. This is in part due to the small sample size included. |

**Supplementary Text, related to Figure 4.**

<u>Modifications to Previously Described Model of AVB-001 Pharmacokinetics</u>

The model used in this analyses was a slightly modified version of a previously developed model for analyzing preclinical data of AVB-001. The original model is given by the following three ordinary differential equations:

$$\frac{dN}{dt} = -\lambda N \tag{S1}$$

$$\frac{dC_P}{dt} = \frac{k_{prod}N}{V_P} - k_{BP}C_P \tag{S2}$$

$$\frac{dC_B}{dt} = k_{BP}\frac{V_P}{V_B}C_P - k_{clear}C_B \tag{S3}$$

This model was modified in two ways. First, equation S2 and S3 was modified to represent the mass of IL-2 in the peritoneal space by substituting $C_P = M_P/V_P$, giving:

$$\frac{dM_P}{dt} = k_{prod}N - k_{BP}M_P \tag{S4}$$

$$\frac{dC_B}{dt} = \frac{k_{BP}}{V_B}M_P - k_{clear}C_B \tag{S5}$$

Additionally, we incorporated body patient body weight (BW) as a covariate influencing volume of the central compartment $V_B$ and $k_{clear}$ according allometric scaling relationships from the literature[1].

$$k_{clear} = k_{clear,0}\left(\frac{BW}{BW_{median}}\right)^{-0.15} \quad \text{(S6)}$$

$$V_B = V_{B,0}\left(\frac{BW}{BW_{median}}\right) \quad \text{(S7)}$$

This model is simulated as an ordinary differential equation with the dose set by the initial condition $N$ based on the amount administered to each patient. Descriptions of each model parameter are found in Supplementary Table S1.

1. Germovsek, E., Cheng, M., and Giragossian, C. (2021). Allometric scaling of therapeutic monoclonal antibodies in preclinical and clinical settings. MAbs *13*, 1964935. 10.1080/19420862.2021.1964935.
